# Single versus repeated intravenous oncolytic reovirus infusions: Implications for immune modulation and rationalised scheduling of therapy in hepatocellular carcinoma

**DOI:** 10.1101/2024.09.26.24314049

**Authors:** Karen J. Scott, Emma West, Rebecca Brownlie, Fay Ismail, Christy Ralph, Matt Coffey, Alan A. Melcher, Alison Taylor, Salvatore Papa, Adel Samson

## Abstract

Scheduling of oncolytic virus (OV) therapy has never been correlated with immunological /clinical response. In hepatocellular carcinoma (HCC) patients, where background liver is frequently chronically injured, repeated dosing may have deleterious implications, resulting in off-target immune-mediated damage, thereby tipping the balance between favourable clinical response and hepatotoxicity. Elucidation of the optimum dosing regime is paramount to ensure therapy, whilst limiting damage to background liver.

We expand upon our experience in neoadjuvant OV therapy to compare immunological responses from single versus repeated doses of reovirus in cancer patients. The impact of OV on HCC outcomes was examined *in vivo* following a high-fat diet or induced liver fibrosis in the context of abnormal background liver. Furthermore, we assess the potential immune-mediated toxicity of single versus multiple virus infusions in combination with PD-1/PD-L1 blockade.

Data indicate that a single dose of reovirus is equivalent or superior to repeated doses in achieving: (a) induction of an inflammatory cytokine/chemokine response; (b) peripheral blood immune cell activation; (c) migration of activated CD8+ CTLs. Repeated doses on consecutive days do not improve the amplitude of immune response following virus infusion. Furthermore, without improving therapeutic efficacy, repeated viral dosing leads to an unwanted influx of activated T-cells into background liver, alongside elevated liver enzymes associated with aberrant liver function. A single dose of reovirus is as effective as multiple doses when combined with anti-PD-L1 therapy in limiting tumour growth and extending survival *in vivo*, whilst simultaneously avoiding undesirable toxicities in background liver, in the context of HCC.

**Novelty and Impact Statement:** In solid malignancies, the optimum dosing schedule of oncolytic virotherapy thus far has not been correlated with clinical or immunological response. In hepatocellular carcinoma, background liver is frequently injured and repeated viral infusions may have harmful repercussions, tipping the balance from favourable clinical response to further hepatotoxicity. Here, we provide pivotal data that elucidates a rationalised reovirus dosing regimen, with and without checkpoint inhibition, which ensures therapy whilst limiting damage to compromised background liver.

## Introduction

Oncolytic viruses (OVs) are a promising immunotherapeutic approach in the treatment of many malignancies, with three agents having been approved for routine therapeutic use^1^. Their mechanism of action is dependent on OV infection and preferential replication in malignant tumour cells with a subsequent ensuing anti-tumoural immune response. OVs exert effects on a patient’s immune system in a multifaceted manner, resulting in many downstream responses. The release of inflammatory factors, including type I IFNs^2,3^, followed by the subsequent induction of a proinflammatory cytokine/chemokine milieu^4–6^, activates innate immune cell populations, both within the tumour and in the peripheral circulation. OV treatment is associated with the elevation of many chemokines necessary for the migration of immune cells^7–10^, including cytotoxic CD8 T-cells, critical mediators of anti-tumour immunity, into areas of inflammation, such as the virus-infected tumour. The migration of lymphocytes from the peripheral blood (PB) in this way results in a transient lymphopaenia^11–13^. Furthermore, infection of tumour cells culminates in tumour cell death, causing release of tumour-associated antigens that are phagocytosed and presented by antigen presenting cells to promote T-cell anti-tumour priming^14^. Although OVs can be administered by intra-tumoural (*i.t*.) injection, the intravenous (*i.v*.) route is frequently preferred due to the convenience of administration and the likelihood of improved virus distribution to metastatic sites of disease. However, despite many finalised OV clinical trials, the optimum scheduling of *i.v*. infusions remains undefined. Trials completed thus far have incorporated both single^15,16^ or multiple consecutive doses^17^ within each two-to four-week treatment cycle, all with the primary aim of maximising OV delivery to the tumour in order to exert its anticancer effect. Furthermore, both individual^17^ and numerous^11,18^ cycles of treatment have been employed.

Whilst many believe that repeated doses within each cycle of treatment are required for therapeutic efficacy, proof of concept for a single shot cure was demonstrated in a patient who received a high intra-arterial dose of measles virus-NIS, where long-term immune control was also observed after therapy^19^. However, evidence for tumour response following single *i.v*. administration is limited in the majority of patients, potentially due to the OVs being depleted by innate immune defences or due to neutralisation by pre-existing anti-viral antibodies^20^. Some evidence suggests that a dose fractionation strategy, i.e. repeated *i.v*. injections of divided doses within each two- to four-week cycle, can produce enhanced intra-tumoural penetration and distribution of OV^21,22^. However, it is not clear whether repeated doses have improved therapeutic value and if they may conversely result in substantially greater side effects. Increased tissue distribution can theoretically lead to inflammation and lymphocytic infiltrates in off-target organs, raising the risk of autoimmune injury. Immune-mediated side effects are particularly important when considering patients with pre-existing chronic organ damage. Chronically damaged organs have less reserve capacity to cope with additional insults, including immune-mediated injury. Furthermore, long-standing organ damage is frequently associated with chronic inflammation, a process that results in the accumulation of abnormal antigens, which are more likely to be recognised as non-self, hence predisposing such organs to immune-mediated injury following OV infusion. Perhaps the best example of this scenario is to be found in patients with hepatocellular carcinoma (HCC), where the majority have underlying liver cirrhosis and reduced hepatic reserves, as a result of chronic liver inflammation, damage and repair. Multiple OVs have been tested in HCC^23^, with mixed results thus far. A key component to the success of OV therapy in HCC is the balance between anti-HCC immunity and off-target immune-mediated damage to the cirrhotic background liver; notably, a specific case reported that administration of a non-replicating adenovirus proved fatal^24^. Rationalised scheduling of *i.v*. infusions is therefore paramount, in order to maximise benefit and minimise harm.

Herein, we build on our experience of neoadjuvant *i.v*. reovirus clinical trials, to compare the PB immune response in patients receiving either a single dose or repeated daily infusions of reovirus for up to five days, within a single cycle of therapy^16,17^. Reovirus is a naturally occurring double stranded RNA OV, which usually results in asymptomatic infection^25^. Oncolytic reovirus has been extensively used in early phase clinical trials and been shown to have a broad range of activity against many malignancies^11,18,26,27^, with studies now in development for its use in the context of HCC. We previously published data from our clinical trials showing successful delivery of virus to tumours following both a single and repeated doses of reovirus^16,17^. Here, we report the nature and amplitude of immunological responses in those patients. Furthermore, using primary tumour cells and PBMCs from HCC patients, as well as HCC cell lines, we show that repeated doses of reovirus have no additional impact on either tumour cell viability or immune activation, over a single dose. Pre-clinical animal models of HCC additionally show that repeated doses of reovirus do not enhance virus delivery to the tumour and are not associated with improved therapeutic efficacy. In contrast, we observed deleterious effects of multiple virus doses on a damaged background liver. Finally, we go on to examine the possibility of combining reovirus therapy with immune checkpoint blockade (ICB) to limit potential further liver toxicities associated with repeated viral dosing.

## Methods

### Reovirus

Clinical-grade Dearing Type III Reolysin® was kindly provided by Oncolytics Biotech Inc., Calgary, Canada. For *in vitro* and *in vivo* studies, reovirus stocks of 2×10^9^ plaque-forming units (pfu)/ml was stored at -80 °C until needed.

### Clinical Trials

#### Reo013 (EUDRACT number 2007-000258-29)

Recruited 10 patients with colorectal cancer metastatic to the liver who were scheduled to undergo planned resection with radical intent. Study recruitment commenced in January 2009 and trial concluded in July 2011. Eligibility/exclusion criteria as previously described^17^. Study patients received 1×10^10^ TCID_50_ Reolysin®, administered daily by *i.v*. infusion over 60 mins, for a maximum of five consecutive days (seven patients received five doses; one patient received four; one patient received three and one patient received one dose). Data from this trial is termed as coming from ‘repeated doses’ of virus.

#### Reo013 Brain (EUDRACT number 2011-005635-10)

Recruited nine patients prior to their planned de-bulking neurosurgery; six with recurrent high-grade glioma and three with metastatic tumours to the brain (one primary colorectal cancer and two primary melanoma). Study recruitment commenced in July 2013 and concluded in November 2014. Eligibility/exclusion criteria as described^16^. Patients received a single infusion of 1×10^10^ TCID_50_ Reolysin®, administered by *i.v*. infusion over 60 mins. Data from this trial is termed as coming from ‘one dose’ of virus.

#### Collection of PB samples from trial patients

PB was collected into K_3_EDTA vacuette tubes (Greiner) and processed, where possible, within one hour of venepuncture. Blood samples were collected as depicted in the trial schemas in Figure 1A.

**Figure 1.**
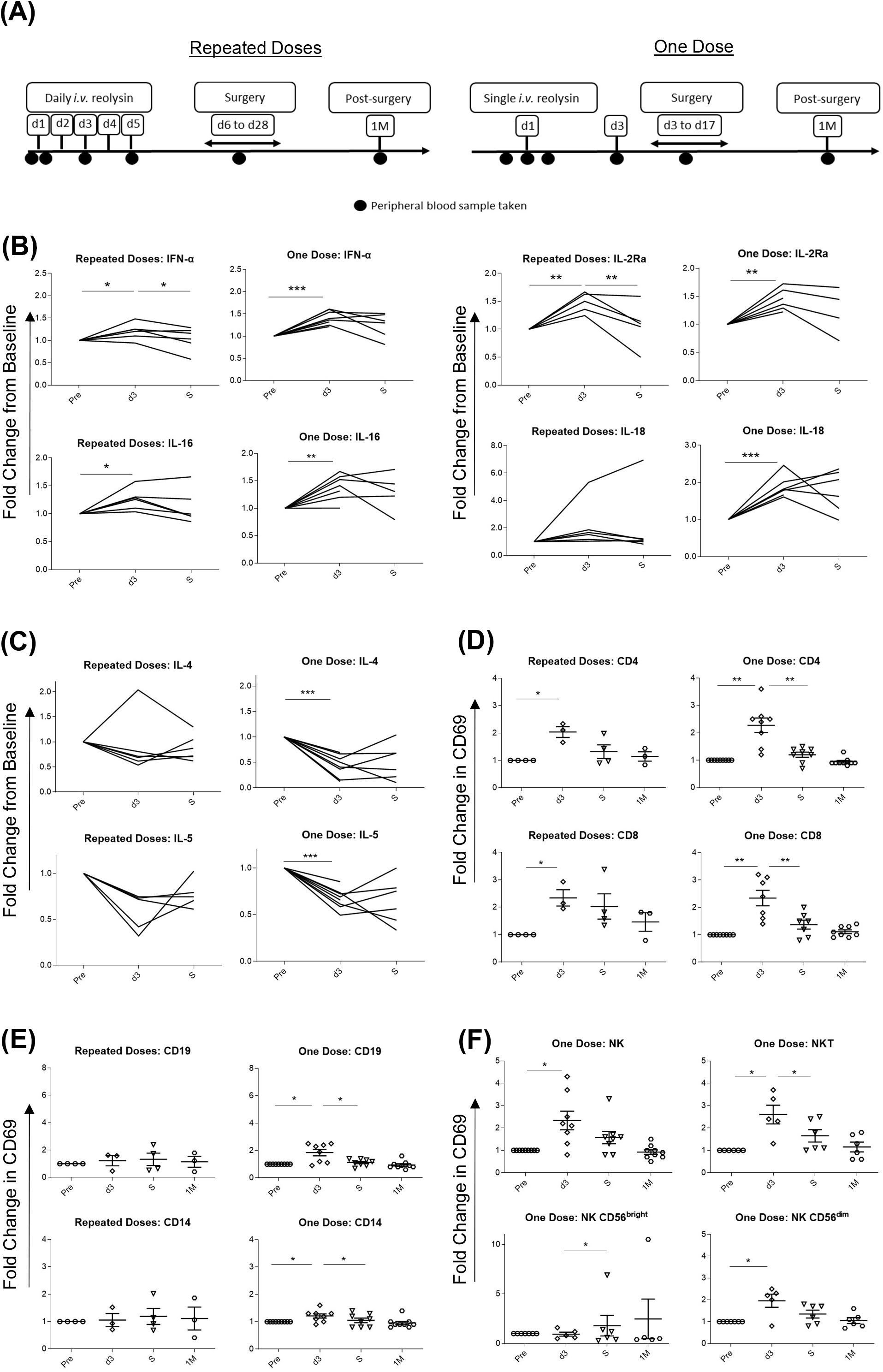
Effects of a single or repeated infusions of *i.v*. reovirus on patient peripheral immunological responses. **(A)** Trial schema for ‘repeated doses’ study and ‘one dose’ study, showing timing of reovirus infusions and PB sample collection. Multi-plex analysis was performed on patient plasma samples taken throughout study periods (Pre-infusion: Pre; day 3: d3; surgery: S; 1 month-post infusion: 1M) to determine secretion of: **(B)** immune-stimulatory and **(C)** immune-inhibitory cytokines. (Data is presented as fold-change from baseline (pre-infusion) sample; each line denotes an individual patient; repeated dose study n=6, single dose study n=9). Whole blood immunophenotyping of patient samples was performed throughout study periods and cell-surface CD69 expression was determined on: **(D)** CD4^+^ T-cells and CD8^+^ T-cells, **(E)** CD19^+^ B-cells and CD14^+^ monocytes and **(F)** CD3^-^ CD56^+^ NK cells, CD3^+^ CD56^+^ NKT-cells, as well as CD56^bright^ and CD56^dim^ NK cells (of those patients given a single dose of reovirus only). (Data is presented as mean ± SEM fold change from baseline (pre-infusion) sample; repeated dose study: n=4; single dose study: n=9. Paired T-tests; *P<0.05, **P<0.01, ***P<0.001).

#### Patient lymphocyte counts

Full blood counts were performed where appropriate as part of standard clinical care. The Patient Pathway Manager and Results Server systems were used to acquire total lymphocyte counts throughout treatment.

#### Isolation of plasma from patient PB

PB was centrifuged for 10 mins at 2000 g and plasma harvested from the resulting upper layer (above red blood cells). Aliquots were stored at –80 °C until required for cytokine/chemokine analysis.

#### Whole blood immunophenotyping of patient samples

Whole blood was added to pre-aliquoted antibodies (see below) and left for 30 mins at 4 °C protected from light. 1X FACS-lysing Solution (BD Biosciences) was added and tubes were immediately vortexed before being incubated for 10 mins at room temperature protected from light. A 300 g spin for five mins at 4 °C was performed, supernatants discarded and FACS buffer (phosphate-buffered saline (PBS) containing 1 % (*v/v*) foetal calf serum (Biosera) and 0.1 % (*w/v*) sodium azide (Sigma)) added before a second spin as above. Cells were fixed in 1 % (*w/v*) paraformaldehyde (Sigma) and stored at 4 °C until flow cytometry was performed. Cells were acquired using a FacsCaliber flow cytometer and analysed using Cell Quest software or an LSR II 3 Laser Cytometer and FACS Diva software (both BD Biosciences). Appropriate isotype controls were used for all immunophenotyping. All post-treatment samples are expressed as relative fold-change compared to pre-treatment samples.

#### Reo013 (repeated doses) antibodies

Anti-human: CD3-APC, CD4-PerCP, CD8-PerCP, CD56-PE, CD14-PerCP, CD19-APC, CD69-FITC (all BD Biosciences).

#### ReoBrain (single dose) antibodies

Anti-human: CD3-PECy7, CD4-V500, CD8-BV500, CD14-BV510, CD19-BV421, CD56-FITC (all BD Biosciences) and CD69-APC (Miltenyi).

#### Chemokine and cytokine multi-plex analysis

Bio-Plex Pro™ Cytokine and Chemokine Assays (21-plex, human group I and 27-plex, human group II; BioRad) were used to determine levels of soluble mediators in plasma samples throughout treatment, as per manufacturer’s instructions. Data is expressed as relative fold change in post-treatment samples compared to pre-treatment samples.

#### Immunophenotyping of HCC patient PBMC

PBMCs were isolated from whole blood, frozen and stored as previously described^28^. Briefly, PBMCs were stained with anti-human: CD3-FITC, CD4-APC-H7, CD8-AF700, CD56-PE-Cy7, CD19-BV421, CD14-PerCP, CD69-APC, PD-L1-PE-CF594 and fixable viability stain-BV510 (all BD Biosciences). Cells were fixed in 1 % (*w/v*) paraformaldehyde and stored at 4 °C until flow cytometry was performed. Fluorescence-minus-one were used as negative controls and data was acquired on a CytoFlex LX cytometer and analysed using CytExpert Software v2.4

#### Cell lines

HLE (RRID:CVCL_1281) and Huh7 (RRID:CVCL_0336) cell lines were purchased from the Japanese Collection of Research Bioresources Cell Bank, have been authenticated using STR profiling and confirmed to be mycoplasma free during the time the experiments were carried out.

#### Reovirus treatment of HCC PBMC, HCC cell lines and primary HCC tumour cells

Cells (1×10^6^/ml) were treated with 1 pfu/cell reovirus or PBS for one or three consecutive days before harvesting on day 4 for subsequent analysis.

#### Cell viability assay

CellTitre-Glo Luminescent cell viability assay (Promega) was used to determine the number of viable cells in HCC cell line or patient-derived single cell tumour cultures, based on quantification of the ATP present as per manufacturer’s instructions.

#### Subcutaneous HCC model

Female BALB/c aged 8-10 weeks were purchased from Charles River Laboratories, UK. To induce tumours, between 1×10^6^ and 1×10^7^ BNL 1 ME A.7R.1 HCC cells (ATCC; RRID: CVCL-6371; routinely tested and found to be negative for mycoplasma infection) were injected subcutaneously (*s.c*.) into the flank and tumour growth was quantified by calliper measurement. Once tumours had established (approximately d21), one or three doses of reovirus or PBS (n=5 or 7 as indicated in figure legend) was administered on consecutive days (d1-3; 1×10^7^ pfu/dose) either *i.v*. or i.t. (as indicated). In some experiments, either 100 µg In Vivo plus anti-mouse PD-L1 (B7-H1) or In Vivo Plus IgG2b isotype control (BioXcell) or 100 µg anti-mouse PD-1 or IgG1 anti-mouse β-Gal isotype control (InvivoFit™, InvivoGen) (as indicated) was administered intraperitoneally (*i.p*.) for three days after reovirus treatment (d4, 5 and 8). This cycle of treatment was repeated at d15. Animal procedures involving subcutaneous HCC tumours were performed under the approved UK Home Office project license PP1816772 (held by AS) in line with the Animal (Scientific Procedures) Act 1986.

#### RT-qPCR for reovirus

Tumours and livers were harvested from *in vivo* experiments and stored in RNAlater (Sigma). Tissues were lysed, homogenised and RNA was extracted using RNeasy Mini Kits (Qiagen). Levels of RNA were quantified using Qubit RNA assay kits and the Quibit 4 Fluorometer (both Thermo Fisher Scientific). cDNA was prepared using the SensiFast cDNA synthesis kit (Meridian Bioscience) and an Applied Biosystems Veriti Thermal Cycler. PCR was performed with targeted primers for reovirus sigma 3 (5⍰-GGGCTGCACATTACCACTGA (forward) and 5’ – CTCCTCGCAATACAACTCGT (reverse; both Sigma) alongside a standard curve of known reovirus concentrations. A detection limit of 35 cycles was used for evaluation on Applied Biosystems QuantStudio™ 5 Real Time PCR Systems (Thermo Fisher). Analysis was performed using the Thermo Fisher Scientific Connect Platform. Data is presented as ΔCT values of reovirus compared to a housekeeping gene, 18s (Sigma).

### Immune infiltration into tumour and liver

The *s.c*. HCC *in vivo* model was initially used to replicate the patient blood sampling schedule. Here, tumours and livers were harvested 72 hours after last virus infusion and stored in formalin for IHC analysis. Formalin-fixed paraffin-embedded (FFPE) tissue sections were stained with rabbit anti-mouse CD4 or CD8 antibody (both Abcam; both 1:500) and ImmPRESS-HRP anti-rabbit IgG (peroxidase) secondary antibody (Vector). Positive staining was visualised using ImmPACT DAB HRP Substrate kit (Vector). Control sections contained no primary antibody. Digital images were acquired at x20 magnification and quantified using Aperio ImageScope software v12.3.3.5048 (Leica Biosystems).

To further investigate the nature and duration of immunological response to reovirus treatment, tumours, livers and tumour-draining lymph nodes (dLN) were harvested at d3 or d9 after the last virus dose and analysed by flow cytometry. Single-cell suspensions were prepared via disaggregation of tissue through a 70 µm cell sieve (Corning) and washes in PBS. Immunophenotyping was performed using anti-mouse CD3-FITC (Miltenyi), CD8-BUV395 (BD Biosciences), CD4-APC-cy7, PD-L1-PE/Dazzle, PD1-BV605, CD69-BV650 (all Biolegend) and analysed by flow cytometry. Data was acquired on a Cytoflex LX cytometer and analysed using FlowJo™ software (BD Biosciences).

#### Liver fibrosis *in vivo* models

Male C57BL/6 mice aged 12 weeks were purchased from Charles River. Mice (n=5/group) were maintained on a high fat diet (HFD) (Fat Calories 60 %, 1/2” Soft Pellets; Bio Serv) for 13 weeks or injected *i.p*. with 0.5 ml/kg body weight of carbon tetrachlorine (CCl_4_; Sigma) diluted in mineral oil twice a week for 4 weeks, followed by once a week for a further 8 weeks. Aged-matched control mice were kept on a normal diet. Groups of mice were then injected *i.v*. with either PBS, one or three doses of 1×10^7^ pfu reovirus on consecutive days. On d4, livers were harvested for analysis by flow cytometry, as described previously. Blood serum (100 µl) was analysed using the 24 Comprehensive Test Plus disc on the SMT-120V fully-automated veterinary biochemistry analyser (Seamaty). Animal procedures involving liver fibrosis were performed under the approved UK Home Office project license PP0972946 (held by SP) in line with the Animal (Scientific Procedures) Act 1986.

#### Statistical analysis

Paired T-tests were performed on clinical trial and HCC patient data to define statistical significance in response to therapy, where indicated. *In vivo* data was analysed using unpaired T-tests and logrank (Mantel-Cox) test. *In vitro* primary and cell line data was analysed using unpaired T-tests. *P<0.05, **P<0.01, ***P<0.001 ****P<0.0001.

## Results

### Reovirus-induced inflammatory cytokine response

To assess the early PB immune response to either a single or multiple *i.v*. infusions of reovirus, we analysed patient blood samples obtained from two separate clinical trials in which patients either received multiple daily infusions (‘repeated doses’) or a single dose (‘one dose’). Patients receiving a single dose were more heavily pre-treated with systemic chemotherapy and radiotherapy and were more likely to be taking steroids (Supplementary Tables 1A and B). Herein, we expand on our previously published clinical observations and show that a significant peripheral immune response to reovirus infusion is achieved by a single dose. PB sample collection is shown in Figure 1A. Plasma samples were analysed for the presence of both cytokines that stimulate cellular anti-cancer immunity (Figure 1B and Supplementary Tables 2A and B) and those that negatively regulate cellular immunity (Figure 1C and Supplementary Tables 2A and B). The results show that regardless of whether a patient is given a single/repeated doses of reovirus, a consistent pattern of cytokine response is detected. In both trials, significantly increased secretion of type I interferon (IFN)-α was observed, as would be anticipated following viral infection^2,3^. Furthermore, many other IFN-stimulated inflammatory cytokines, for example, IL-2Ra, IL-16 and IL-18, were also elevated by d3 after virus infusion regardless of whether patients had received a single/repeat infusions. Conversely, Th2 cytokines, IL-4 and IL-5, decreased following reovirus treatment in both patient groups, amounting to a significant reduction in concentrations following a single but not repeated infusion. The peak in proinflammatory cytokines and the recovery of IL-4 and IL-5 return to pre-treatment levels by the time of surgery regardless of the number of infusions received. Overall, this indicates that both single and repeated doses of virus have similar effects in the induction of a pro-inflammatory (Th1) systemic immune environment, in conjunction with a dampening of the anti-cellular immunity (Th2) cytokine response, suggesting that multiple doses are not necessary in the initial stages of immune anti-tumour activation, following reovirus infusion.

### Reovirus-induced immune cell activation

As may be predicted by the induction of a pro-inflammatory cytokine response, both repeated/single doses of reovirus caused similar activation of CD4+ and CD8+ T-cells, demonstrated by an upregulation in expression of CD69, an early activation marker (Figure 1D). CD69, known to be upregulated in response to many cytokines, including type I IFNs and IL-18^29–31^, increased significantly by d3, before returning to pre-treatment levels by surgery following both repeated/single doses of reovirus. Very early changes in CD69 expression, either during virus infusion or 60 minutes post-infusion, were not observed (Supplementary Figure S1 and Supplementary Tables 3A and B). A similar peak at d3 was observed on CD19+ B-cells and CD14+ monocytes, in response to a single infusion, although not in response to repeat infusions (Figure 1E and Supplementary Tables 3A and B). Furthermore, in patients receiving a single dose of reovirus, an elevation in CD69 expression on both NK (specifically cytotoxic CD56^dim^ cells) and NKT cells was also apparent (Figure 1F). Whilst NK and NKT cell CD69 expression wasn’t assessed on patients receiving repeat infusions, these data again demonstrate that a single dose, within any given cycle of reovirus treatment, is sufficient for the activation of multiple immune sub-sets, with the peak of activation occurring at d3.

### Reovirus-induced fluctuations in peripheral T-cells

Analysis of patients’ full blood counts during reovirus therapy revealed a transient lymphopenia at d3, similar in magnitude following both single/repeated infusions, which returned to baseline levels in subsequent weeks (Figure 2A). Flow cytometric analysis of specific immune cell subtypes revealed that many lymphocytes decreased proportionally in the PB, contributing to this lymphopenia; CD4+ T-cells are at a significantly reduced frequency following both single/repeat doses of reovirus, whilst CD8+ T-cells were significantly reduced following a single, but not repeated doses (Figure 2B). Analogous to this, CD19+ B-cells are also present at lower levels on d3, again more marked following a single rather than repeated doses (Figure 2C). In contrast, the proportion of CD14+ monocytes appears elevated in PB after viral therapy at d3, rising proportionally to other lymphocyte subsets (Figure 2C). In keeping with the observed lymphopenia, changes in the concentrations of many chemokines that are associated with the trafficking of immune cells were apparent when patient plasma was examined. Specifically, interferon gamma-induced protein 10 (IP-10), monokine induced by gamma-interferon (MIG) and macrophage inflammatory protein 1-β (MIP-1β) levels peaked at d3 following virus infusion, before falling back to baseline at subsequent time points (Figure 2D). These data indicate that a single reovirus infusion is sufficient or potentially superior to multiple infusions in creating an immune environment in the blood, which may be permissive to lymphocyte migration into the tumour. Repeated doses predominantly do not amplify or prolong cytokine/chemokine secretion significantly over a single dose (Supplementary Figure S1 and Supplementary Tables 2A and B, 4A and B). Notably, this equivalent response to a single dose compared to repeated doses occurred in the patients who were more heavily pre-treated with chemo/radiotherapy and more likely to be receiving steroids at the point of reovirus infusion.

**Figure 2.**
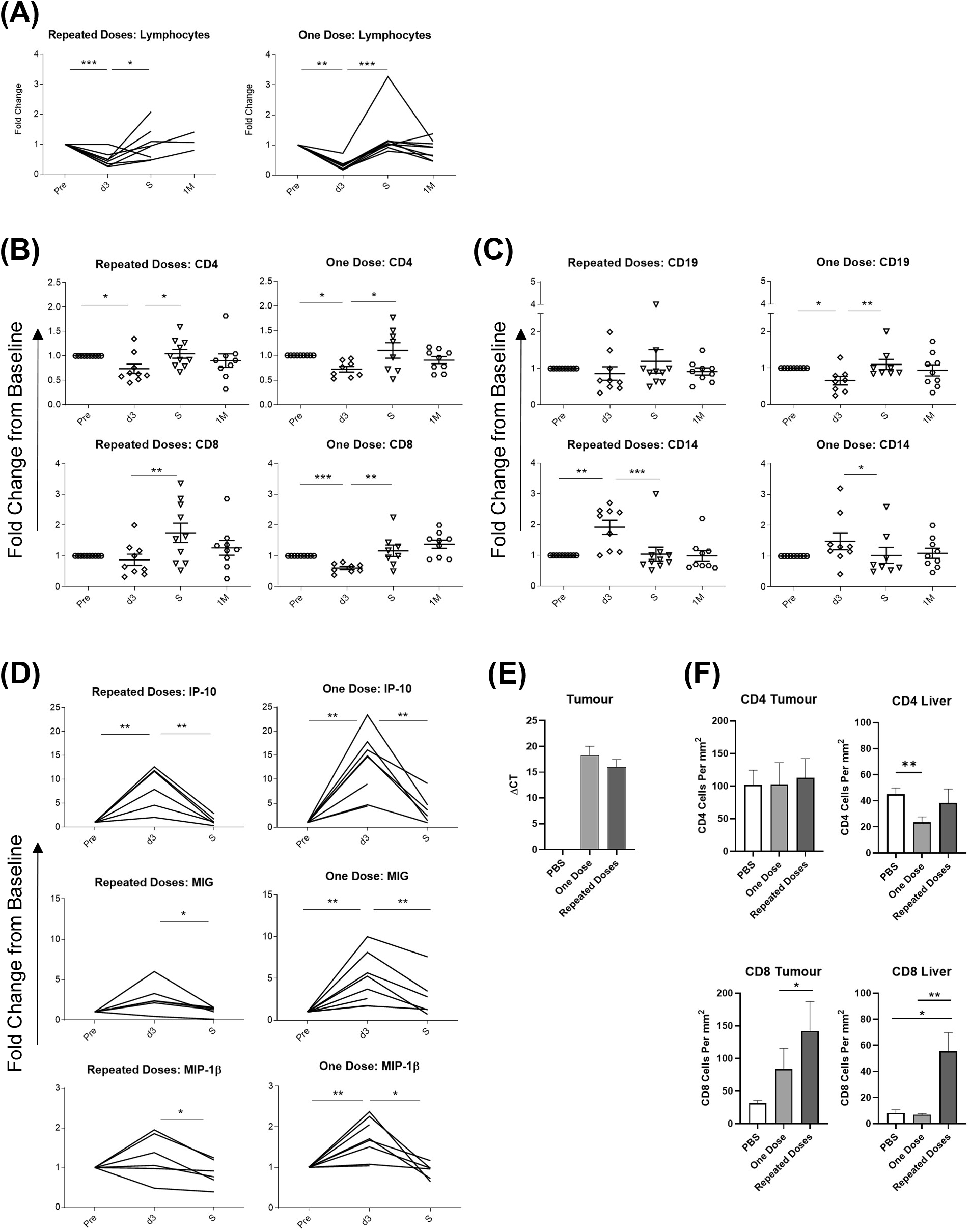
Effect of single or repeated infusions of *i.v*. reovirus on relative abundance of T-cells in PB and tissue. **(A)** Changes in patient total lymphocyte count over the treatment period (Pre-infusion: Pre; day 3: d3; surgery: S; 1 month-post infusion: 1M). (Data is presented as fold-change from baseline (pre-infusion) with each line representing an individual patient). Whole blood immunophenotyping of patient samples was performed to determine changes in: **(B)** CD4^+^ T-cells, CD8^+^ T-cells and **(C)** CD19^+^ B-cells and CD14^+^ monocytes. (Data is presented as mean ± SEM fold-change from baseline (pre-infusion) sample; repeated dose study n=10, single dose study n=9). **(D)** Multi-plex analysis was performed on patient plasma samples taken throughout study periods and changes in secretion of the chemokines IP-10, MIG and MIP-1β were determined. (Data is presented as fold change from baseline (pre-infusion) sample; each line represents an individual patient; Repeated dose study n=6, single dose study n=9; Paired T-tests; *P<0.05, **P<0.01, ***P<0.001). 1MEA tumour-bearing mice were treated with PBS control (white bars) or by a single (light grey bars) or repeated (dark grey bars) *i.v*. doses of reovirus. **(E)** RT-PCR was performed on tumours for the presence of reovirus sigma 3 and is shown as ΔCT of reovirus compared to the 18S housekeeping gene. **(F)** IHC was performed on FFPE tissue for CD4+ and CD8+ T-cells and quantified as immune cell positivity per mm^2^ of tissue. (Data is presented as mean ± SEM for n=5 per group; unpaired T-tests; *P<0.05, **P<0.01).

### Reovirus-induced T-cell infiltration in HCC

The need to balance clinical efficacy and toxicity with intravenous OV therapy is perhaps best illustrated in patients with HCC, where the background liver is highly susceptible to immune-mediated damage. To determine the potential effectiveness of single versus repeat reovirus doses in HCC, we utilised a syngeneic immunocompetent model in Balb/c mice. Mice were administered with either one or repeated doses of *i.v*. reovirus to determine the pre-clinical effect on both virus delivery and T-cell infiltration into tumour and liver. Administering repeated doses of reovirus to tumour-bearing mice did not result in greater targeting of virus to tumour tissue (Figure 2E), indicating no therapeutic advantage of multiple doses over a single dose; reovirus was cleared from the livers by the time of organ harvest (data not shown). Figure 2F and Supplementary Figure S2 depict the influx of CD4+ and CD8+ T-cells into the tumour and liver at d3 following one dose or the last of repeated doses of reovirus. Interestingly, the level of CD4 T-cells within the tumour are not increased by reovirus; in fact, one dose causes a reduction in CD4 T-cells within the liver. In contrast, there is a significant increase in CD8+ staining within the tumour and an even greater CD8+ T-cell infiltration into the liver, which, in the context of a damaged liver, may result in erroneous immune-mediated deleterious effects.

### Potential off-target effects of reovirus therapy on chronically damaged HCC livers

As previous data indicates a considerably greater influx of CD8+ T-cells into the liver following reovirus treatment, we examined the toxicity of reovirus therapy on chronically damaged livers in order to model the scenario in HCC patients with impaired background liver function. Liver damage was induced in mice before administering either single or multiple doses of reovirus, followed by assessment of liver damage and immune cell infiltration. One cohort was maintained on a HFD for 13 weeks, gaining on average over 90 % of their starting weight (Supplementary Figure S3), while another cohort was injected with CCl_4_ to induce hepatic fibrosis. After reovirus delivery, liver T-cells were assessed by flow cytometry for CD69 and PD-L1 expression whilst biochemistry analysis was performed on serum to measure liver enzyme activity, including aspartate aminotransferase (AST), alanine transaminase (ALT), albumin (ALB), total bile acids (TBA), alkaline phosphatase (ALP) and creatine kinase (CK) (Figure 3). Within the cohorts, administration of either a single/multiple doses of reovirus resulted in a similar increase in expression of CD69 and PD-L1 on hepatic-resident CD4 and CD8 T-cells, showing no further T-cell activation advantage of multiple doses (Figure 3A). With regard to liver function (Figure 3B), AST, ALT, ALB and ALP from control mice injected with PBS all fell within the normal range of detection (indicated by the grey band). Reovirus did not significantly alter this, with the exception of ALB, which was comparably reduced by both single/multiple doses of reovirus, a pattern seen across all cohorts of mice; this is indicative of reduced liver function. Similarly, CCl_4_ injections alone had marginal effect on enzyme levels; however, the administration of reovirus to these mice resulted in elevations of AST and ALT to above that of the healthy range, again suggesting liver cell lysis and reduced liver function, respectively. No significant differences in levels of any solutes between single/multiple doses were observed in CCl_4_-pretreated mice. Mice fed the HFD and treated with PBS had high levels of AST and ALT compared to control mice. A single dose of reovirus had no effect over and above PBS for both AST and ALT in HFD-mice, yet AST was significantly increased (two-fold over a single reovirus dose and over five times the healthy range) upon multiple reovirus injections, suggesting a greater degree of liver cell lysis. The variable, albeit largely unchanged, levels of CK suggest that this AST increase was a result of liver and not myocyte damage. Furthermore, multiple doses of reovirus also increased TBA in CCl_4_ and HFD mice and, in some cases, ALP in HFD mice. Overall, hepatic T-cell activation by single/multiple doses of reovirus is comparable; however, in the context of a damaged liver, multiple doses are potentially more harmful than a single dose, as indicated by a change in solutes which are associated with a pronounced reduction in liver function.

**Figure 3.**
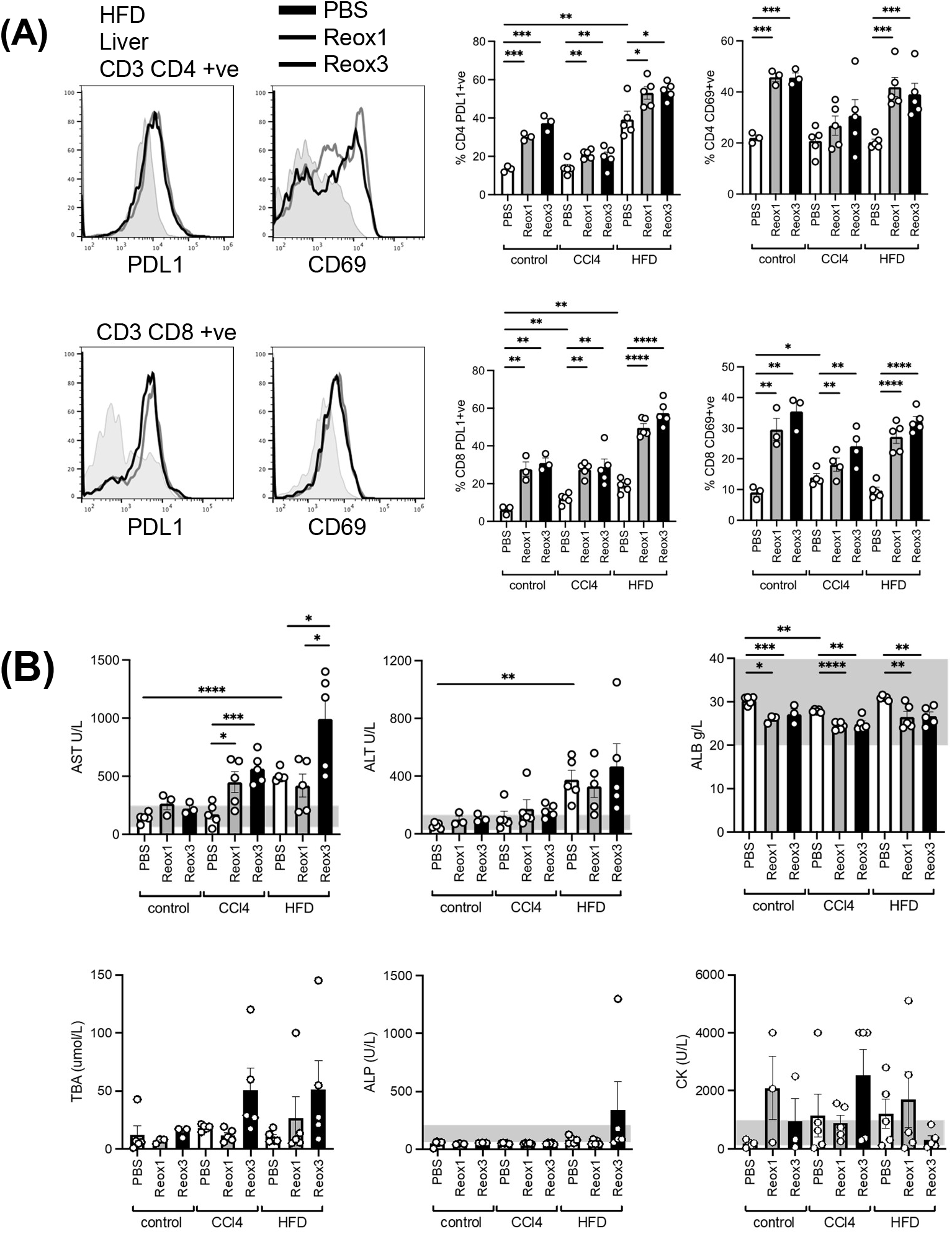
Effect of single or repeated infusions of *i.v*. reovirus on a physiologically-damaged background liver. PBS (white bars), a single dose (Reox1; grey bars) or repeated doses (Reox3; black bars) of reovirus were administered to mice with induced liver damage by HFD or CCl _4_, or to control mice, by *i.v*. injection before harvest of livers on d4. **(A)** Flow cytometry analysis of liver samples for PD-L1 and CD69 expression on CD4+ and CD8+ T-cells are depicted. Representative histograms and bar charts presented as mean ± SEM % positive cells. **(B)** Liver function was assessed by measuring serum levels of AST, ALT, ALB, TBA, ALP and CK. Grey boxes represent the normal range. (Data is presented as mean ± SEM solute concentration; n=5 per group; unpaired T-tests *P<0.05, **P<0.01, ***P<0.001, ****P<0.0001).

### Efficacy of reovirus can be enhanced by combination with PD-L1

Given our previous data showing that a single dose of reovirus was sufficient to activate T-cells (Figures 1, 2 and 3A), we sought to determine the expression profiles of reovirus-induced PD-1 and PD-L1 on T-cells in tumour-bearing mice after a single dose. Figure 4 shows that PD-L1 was significantly upregulated on a large proportion of CD4 and CD8 T-cells in dLN (>90 %), liver (>80 %) and tumour (>60 %) 3 days after *i.v*. reovirus, with levels reducing by d9. Conversely, reovirus did not upregulate PD-1 expression on CD8 T-cells and only a small proportion (<5 %) of dLN and liver CD4 T-cells by d9 (Figure 4A and B). CD69 was also observed to increase following a single reovirus injection by d3, most notably on dLN T-cells. Similarly, HCC patient PBMCs also induced an equivalent upregulation of CD69 and PD-L1 on CD4+, CD8+ T-cells (Figure 4C), NK cells, CD14+ monocytes and CD19+ B-cells (Supplementary Figure S4) following exposure to reovirus, whilst repeated doses had limited additional effect over one dose.

**Figure 4.**
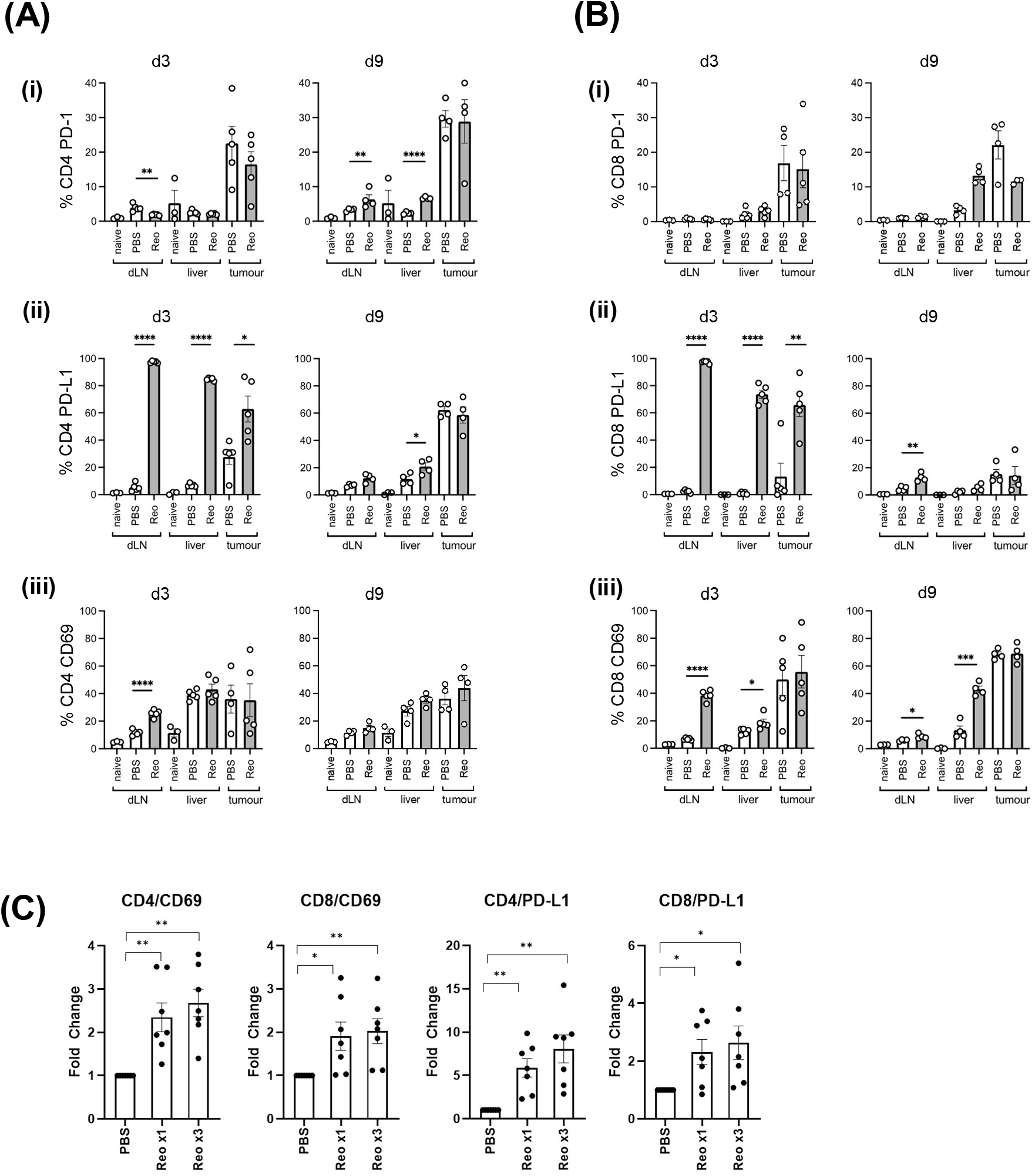
Effects of single or repeated doses of reovirus on checkpoint molecule and CD69 expression on murine T-cells in dLN, liver and tumour and HCC patient PBMC. 1MEA tumour-bearing mice were treated with a single dose of *i.v*. reovirus or PBS (naïve represents untreated mice) prior to organ harvest at 3 or 9 days post-treatment. Single cell suspensions of dLN, liver and tumour were analysed by flow cytometry for: **(i)** PD-1, **(ii)** PD-L1 and **(iii)** CD69 expression on **(A)** CD4 and **(B)** CD8 T-cells. (Data is presented as mean ± SEM % positive cells for naïve, PBS (white bars) and reovirus (grey bars); n=5; unpaired T-tests *P<0.05, **P<0.01, ***P<0.001). **(C)** PBMCs from HCC patients were treated with PBS, a single dose (Reox1) or multiple doses (Reox3) of reovirus prior to assessing CD69 and PD-L1 on CD4+ and CD8+ T-cells (individual patient data (n=7) is shown ± SEM; paired T-tests *P<0.05, **P<0.01).

We further sought to define whether *i.t*. injection would enable more focused tumour immune activation, whilst protecting the background liver. Although *i.t*. injection appeared to upregulate CD69 expression to a greater level than the *i.v*. route, predominantly in tumour compared to liver, no significant differences in PD-1 and PD-L1 upregulation were observed between *i.v* and *i.t* administration (Supplementary Figure S5).

We next determined the potential for combination reovirus-PD-1/PD-L1 blockade in HCC, in the context of multiple/single reovirus injections (Figure 5). As expected, both single/multiple doses of reovirus alone had no impact on tumour burden or survival. In contrast, *in vitro* data demonstrated a negative effect of reovirus on the metabolic activity of both HCC patient-derived tumour cells and HCC cell lines to an equivalent level, regardless of the number of reovirus doses given (Supplementary Figure S6). Combining single/multiple injections of reovirus with anti-PD-1 also did not significantly change tumour growth or survival (Figures 5A & B). However, the addition of anti-PD-L1 in combination with reovirus did significantly delay tumour growth and prolong survival (Figures 5C & D). Importantly, multiple doses of reovirus did not enhance the efficacy of this combination therapy over that observed from a single dose. These data suggest that an effective combination of reovirus plus anti-PD-L1 could potentially be achieved with a single dose of virus within each cycle of therapy.

**Figure 5.**
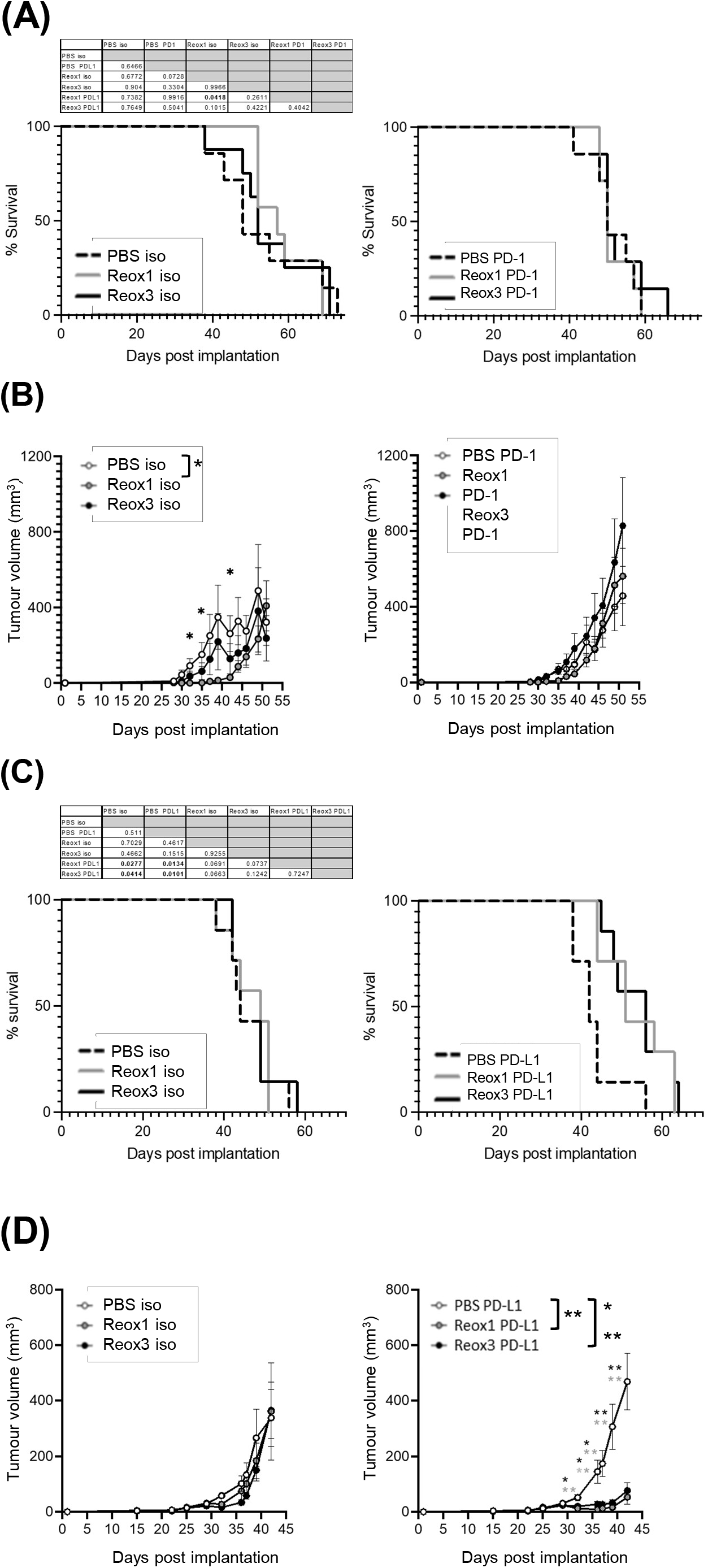
A single dose of reovirus in combination with PD-L1, rather than PD-1 blockade, delays tumour growth. 1MEA tumour-bearing mice were treated with PBS (dashed line /white circles), a single (Reox1; grey) or repeated (Reox3; black) doses of *i.v*. reovirus followed by three consecutive days of *i.p*. anti-PD-1 or anti PD-L1 or respective isotype controls. This treatment regime was repeated again 15 days after first treatment began. Survival curves (**A** and **C**) and tumour growth (**B** and **D**) are depicted for combination with PD-1 or PD-L1, respectively. (Tumour growth curves are shown as mean ± SEM tumour volume (mm^2^) for n=7 per group; unpaired T-tests *P<0.05, **P<0.01).

## Discussion

Despite a plethora of both pre-clinical and clinical trials over recent years investigating the potential of OVs in the immunotherapeutic treatment of many cancer types, the optimum scheduling and dosing regime on patient immune responses has not yet been elucidated. A single ‘one-shot’ dose avoids the subsequent neutralisation by virus-specific antibodies, although a multiple-dose approach may, in theory, be more immunologically or clinically effective due to greater exposure to virus. To date, the scheduling of OV therapy, with single versus multiple infusions on consecutive days, has not been correlated with immunological or clinical response in any solid malignancies. HCC tumours have a considerably more problematic physiology; chronic injury to the liver due to lifestyle factors or HBV/HCV infection induces long-term damage, which is highly susceptible to off-target immune-mediated harm. In these patients, repetitive dosing may have undesirable effects to the damaged liver, resulting in hepatotoxicity, which may outweigh the clinical advantages of therapy.

Although enhanced delivery of reovirus to the tumour might be anticipated with repeated doses, particularly following the onset of immune exhaustion after the first dose, we have previously shown that a single dose is able to penetrate and infect tumour tissue^16^. Furthermore, as OVs replicate in malignant cells, the delivery of a lower dose to tumour may be sufficient for subsequent viral amplification, bypassing the need for repeated doses. Herein, we show that a single dose is sufficient to deliver an equivalent level of virus to the tumour as with repeated doses, meaning additional treatments are not necessary for therapeutic efficacy. In addition, despite a proportionally negative impact on the viability of both primary HCC tumour cells and HCC cell lines following single or repeated doses of reovirus, there was no association with therapeutic efficacy in the *in vivo* model as a single agent.

We further present data in an amalgamated study from two separate clinical trials using reovirus in different dosing regimens that indicates a single dose is adequate to achieve immune activation and may, therefore, be sufficient to generate a subsequent tumour-specific pro-inflammatory environment, equivalent to that generated by multiple doses. Our data show a single dose results in PB elevations in cytokines, such as IL-2Ra, IL-16 & IL-18, which are involved in stimulating T-cells^4,5^ and NK cells^6^ (critical cells in the anti-cancer response following OV therapy) alongside reductions in Th2 cytokines known to be involved in suppression of cellular anti-cancer immunity (IL-4 & IL-5)^32^. Furthermore, greater plasma concentrations of chemokines primarily implicated in immune cell chemotaxis and tissue infiltration of CD8+ T-cells^7^, CD4 T-cells^8^, NK cells^9^ and monocytes^10^ were evident following single/multiple infusions. IP-10 & MIG are predominantly implicated in CD8+ T-cell recruitment^33^ and are likely associated with the significantly greater influx of CD8+ T-cells into tumour and liver in our *in vivo* models, which was not observed with CD4+ T-cells. Overall, this indicates that a single infusion of oncolytic reovirus induces a comparable and possibly improved PB inflammatory cytokine and chemokine response in comparison to repeated consecutive infusions of virus in patients. Although observed in distinct tumour types, both patient groups studied were comparable with regard to baseline clinical factors (Supplementary Tables 1A and B); whilst this may limit observations on clinical efficacy, we have used these two patient cohorts as a representative tool to demonstrate the immunological effects of reovirus infusion. Furthermore, the ‘one dose’ cohort was heavily pre-treated with both chemotherapy and radiotherapy, whilst also predominantly on steroid therapy prior to and throughout the treatment and blood sampling period, in contrast to the multiple dose cohort of patients. Despite the highly immunosuppressive properties of these treatments, patients in the single dose cohort still mounted a significant immunological response to reovirus, which was equivalent to, or greater than, patients receiving additional reovirus infusions in the absence of such treatments. Furthermore, additional studies using PBMCs isolated from the PB of HCC patients showed similar increases in immune cell activation regardless of the number of treatment doses, demonstrating the immune-modulatory effect is applicable in HCC.

A single infusion was also associated with equivalent, if not greater, PB lymphopenia, which is commonly observed following both pathogenic and therapeutic virus infection^11–13^ and is associated with immune cell activation and migration of immune cells away from the vascular compartment, likely to sites of virus infection in tumour, where an adaptive response can be initiated. The fact that multiple virus infusions had no marked enhancement over a single dose likely represents immune cell refractoriness following the initial stimulation by the first dose, a previously observed phenomenon^34^. Likewise, repeated dosing regimens have been associated with diminishing results with each successive OV treatment^35^. In fact, there are no preclinical or clinical studies which clearly demonstrate the superiority of repeat administrations over a single dose within each cycle of treatment^35^. Utilising complementary *in vivo* studies, we show, in parallel to the evidence that a single dose is immunologically equivalent to multiple doses in patients, that unfavourable responses are associated with repeated infusions; a greater influx of proinflammatory T-cells into the liver following multiple reovirus doses which, in the context of HCC, may have damaging off-target effects.

Almost all HCC patients have underlying liver fibrosis or cirrhosis^36^; the most common aetiologies being non-alcoholic fatty liver disease, non-alcoholic steatohepatitis, autoimmune hepatitis, metabolic syndrome, viral hepatitis and alcohol consumption^37^, all of which are strong risk factors for HCC. To assess the impact of liver health on the ability to tolerate reovirus, we used two models of chronic liver damage: HFD and multiple injections of CCl_4_ to induce liver fibrosis. In both settings and in keeping with our clinical trial data, the activation of T-cells by a single dose was comparable to multiple doses of reovirus. However, in general, mice subjected to liver damage, particularly HFD, had abnormal amounts of liver enzymes, which were further exacerbated by reovirus administration. Most strikingly, AST was increased up to five times the healthy range when reovirus was given in multiple doses to HFD mice in comparison to a single dose or PBS. Patients with greater than five times the upper limit of normal AST are considered to have moderate hepatocellular injury^38^. Given that reovirus was cleared from murine livers within a short time, the fact that deleterious effects were still being detected suggest a prolonged impact on damaged background livers long after any therapeutic benefit, with additional doses posing a greater risk. Taken together, these studies highlight the need to carefully balance the potential damaging effects of multiple doses of reovirus with beneficial clinical outcome in the context of HCC.

Previous studies have shown no clinical benefit of reovirus monotherapy (reviewed in^39^). It is now recognised, however, that OV monotherapy induces expression of the immune checkpoint receptor, PD-1 and its ligand PD-L1, which act to limit immune-mediated injury but also inhibit the efficacy of OVs^16,40,41^. A multi-modal combination approach is therefore necessary to optimise OV effectiveness. Multiple early-phase clinical trials employing the combination of an OV with PD-1/PD-L1 blockade have shown good evidence for synergistic efficacy^16,40,42–44^. More recently, PD-L1 inhibition has become standard of care in advanced HCC^45^, usually in combination with anti-vascular endothelial growth factor (VEGF) or anti-cytotoxic T-lymphocyte antigen 4 (CTLA-4) inhibition, but also as Durvalumab monotherapy^45,46^. Our HCC *in vivo* model demonstrates a highly significant transient elevation in PD-L1 expression on both CD4+ and CD8+ T-cells from tumours, livers and dLNs in response to a single reovirus injection, peaking at d3 then reducing at later time points. This transient upregulation was utilised in a combinatorial approach in an *in vivo* model of HCC. Here, we show reovirus alone has no impact on tumour burden, yet when combined with anti-PD-L1 antibodies, reovirus could significantly delay tumour growth and prolong survival. Although initial investigations indicated that repeated doses were associated with a greater influx of CD8+ T-cells into tumour, this did not result in improved efficacy in combination with anti-PD-L1 therapy, meaning that a single dose was as effective as multiple doses. The same phenomenon was not observed when using anti-PD-1/reovirus combination. Similarly, phase III clinical trials have demonstrated that anti-PD-L1 therapy was significantly effective in HCC^46,47^, in contrast to anti-PD-1^48^. These data suggest that an effective combination reovirus/PD-L1 approach would only require a single dose of virus within each cycle of therapy, thereby limiting potential further damage to an already compromised background liver. This merits further investigation in a larger patient group with earlier stage tumours, who are more likely to respond in a clinically beneficial way.

In HCC, strategies for combination immunotherapies, such as OV with ICB, are currently being tested in clinical trials (NCT04665362). In the context of HCC, however, the background liver tissue is usually abnormal or cirrhotic and particularly susceptible to immune-mediated injury. This is exemplified by the higher rates of hepatic injury following ICB in patients with HCC, in comparison to other cancer types^49–51^. Studies have shown that ICB is associated with the infiltration of large numbers of CD8+ T-cells into background liver tissue^52^. Hence, the combination of OV and ICB therapy should be scheduled to limit the infiltration of immune cells into background liver tissue, to reduce the incidence and severity of immune-mediated hepatitis, particularly in the treatment of HCC. Our data collectively suggest that a combinatorial approach comprising a single reovirus infusion within each cycle of therapy is likely to be safest for the background liver and equally as effective in inducing anti-tumour immunity.

## Data Availability

All data produced in the present study are available upon reasonable request to the authors

## Abbreviations

ALB: Albumin
ALP: Alkaline phosphatase
ALT: Alanine transaminase
AST: Aspartate aminotransferase
CCl_4_: Carbon Tetrachloride
CK: Creatine kinase
CTLA-4: Cytotoxic T-lymphocyte antigen 4
dLN: tumour-draining lymph node
FFPE: Formalin-fixed paraffin-embedded
HCC: Hepatocellular carcinoma
HFD: High-fat diet
ICB: Immune checkpoint blockade IFN Interferon
i.p.: intra-peritoneal
IP-10: Interferon gamma-induced protein 10
i.t.: Intra-tumoural
i.v.: Intravenous
MHRA: Medicines and Healthcare Products Regulatory Authority
MIG: Monokine-induced interferon gamma
MIP: Macrophage inflammatory protein
OV: Oncolytic virus
PB: Peripheral blood
PBS: Phosphate buffered saline
s.c.: Subcutaneous
TBA: Total bile acids
VEGF: Vascular endothelial growth factor

## Additional Information

## Acknowledgements

We are grateful to all the patients who participated in these trials

## Author Contributions

Karen J. Scott, Emma West and Rebecca Brownlie: substantial contributions to the conceptualisation, methodology, investigation, formal analysis, visualisation, writing the original draft and review/editing of the final manuscript. Fay Ismail and Alison Taylor: contribution to the investigation and review of the final manuscript. Christy Ralph: contribution to the review of the final manuscript. Matt Coffey and Alan A. Melcher: substantial contributions to the conceptualisation and review/editing of the final manuscript. Salvatore Papa: substantial contribution to the conceptualisation, resources and review/editing of the final manuscript. Adel Samson: substantial contribution to the conceptualisation, methodology, resources, supervision, funding acquisition, writing the original draft and review/editing of the final manuscript.

## Ethics Statement

### Statement of Ethics

Both clinical trials were conducted at The Leeds Teaching Hospitals NHS Trust (LTHT), Leeds, UK and performed in accordance with the Declaration of Helsinki and Good Clinical Practice guidelines (GCP).

### Consent to participate statement

All patients gave written informed consent according to GCP guidelines.

### Study approval statement and clinical trial registration

Reo013 ‘repeated doses’ study; EudraCT number 2007-000258-29. This study protocol was reviewed by the following committees and given the relevant approval numbers: REC reference 08/H1306/73; LTHT R&D approval number CO06/8048. Reo013 Brain ‘single dose’ study; EudraCT number 2011-005635-10. This study protocol was reviewed by the following committees and given the relevant approval numbers: REC reference 18/LO/0080; IRAS project ID 235809.

### Data Availability Statement

All data generated or analysed during this study are included in this article and its supplementary material files. Further enquiries can be directed to the corresponding author.

