## Supplementary Figure 1 for "Single versus repeated intravenous oncolytic reovirus infusions: Implications for immune modulation and rationalised scheduling of therapy in hepatocellular carcinoma"

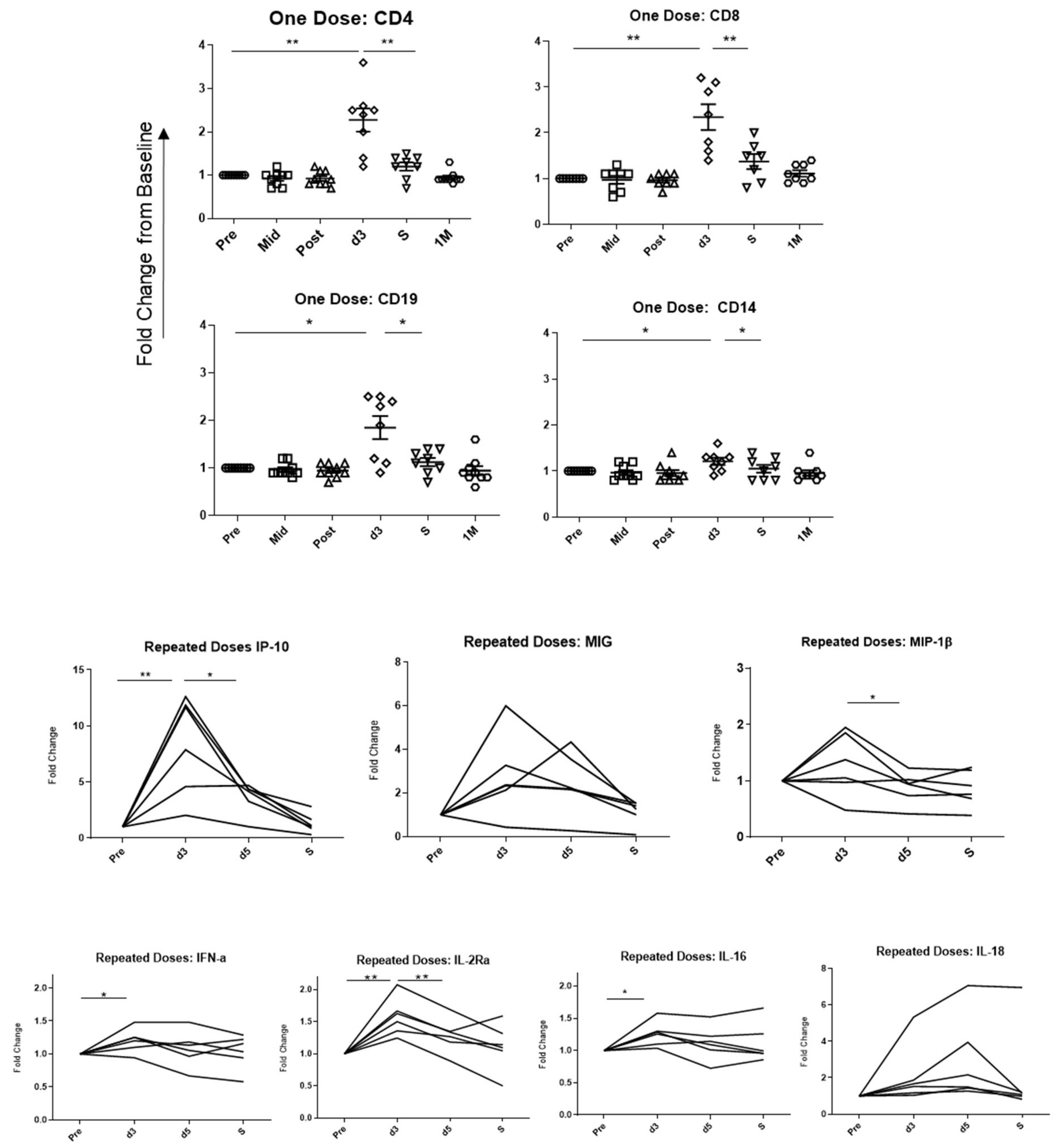


**<u>Supplementary Figure S1: Time course of activation of PB immune cells and chemokine/cytokine secretion in response to single or repeated doses of *i.v.* reovirus.</u>**

Whole blood immunophenotyping for cell-surface CD69 and multi-plex solute analysis on plasma was performed throughout study periods (including mid-infusion, 1hr post-infusion and day 5 samples).

(Data is presented as fold change from baseline (pre-infusion) sample; *P<0.05, **P<0.01. Repeated dose study: n=6; single dose study: n=9.)
