## Supplementary Figure 2 for "Single versus repeated intravenous oncolytic reovirus infusions: Implications for immune modulation and rationalised scheduling of therapy in hepatocellular carcinoma"

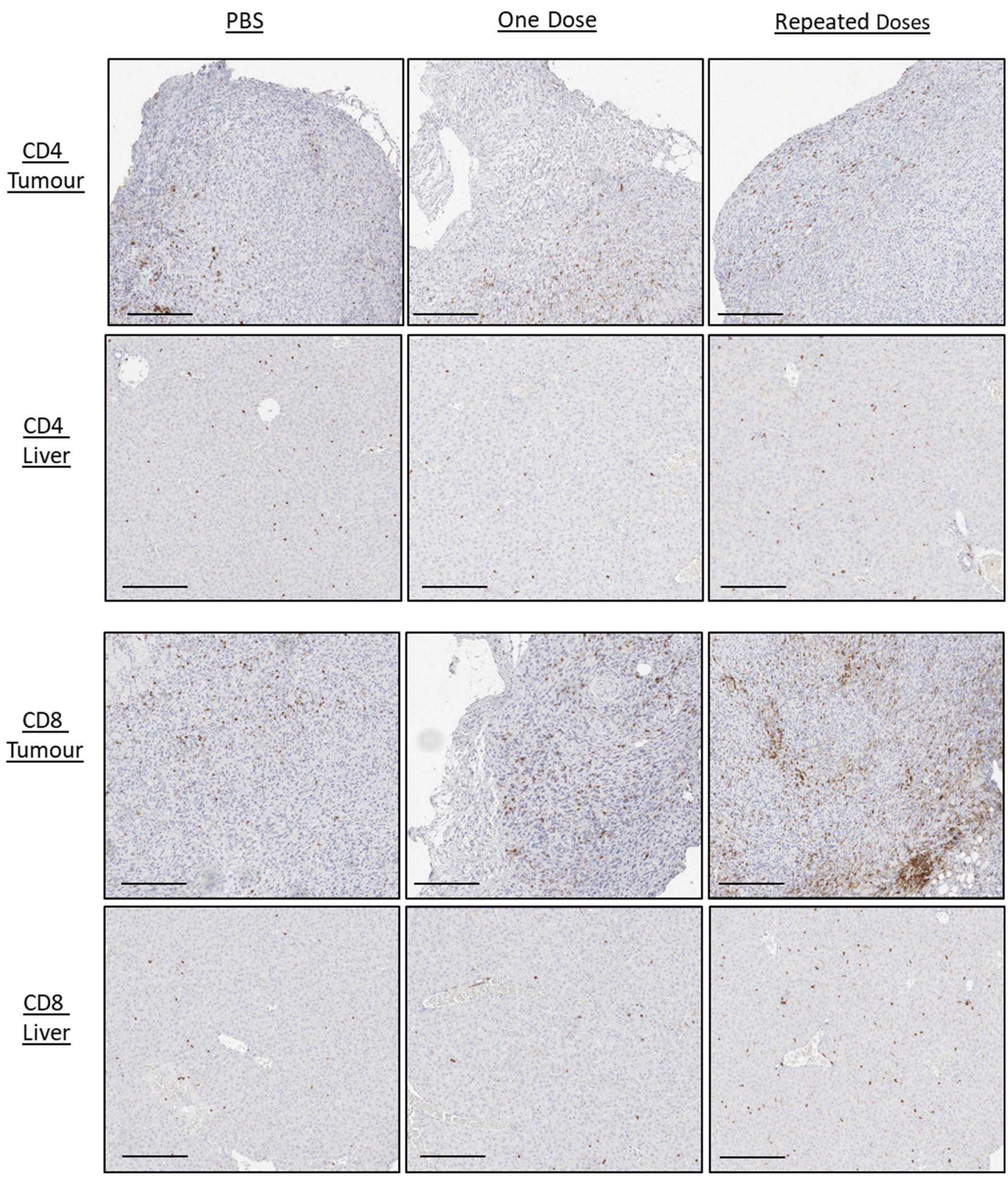


**Supplementary Figure S2:** 1MEA tumour-bearing mice were treated *i.v.* with PBS, a single or repeated doses of reovirus. Livers and tumours were harvested 72 hours after the final virus dose. IHC was performed on FFPE tissue for CD4+ and CD8+ T cells. Representative images are shown; positive staining is DAB (brown) and scale bars represent 100 µm.
