## Supplementary Figure 3 for "Single versus repeated intravenous oncolytic reovirus infusions: Implications for immune modulation and rationalised scheduling of therapy in hepatocellular carcinoma"

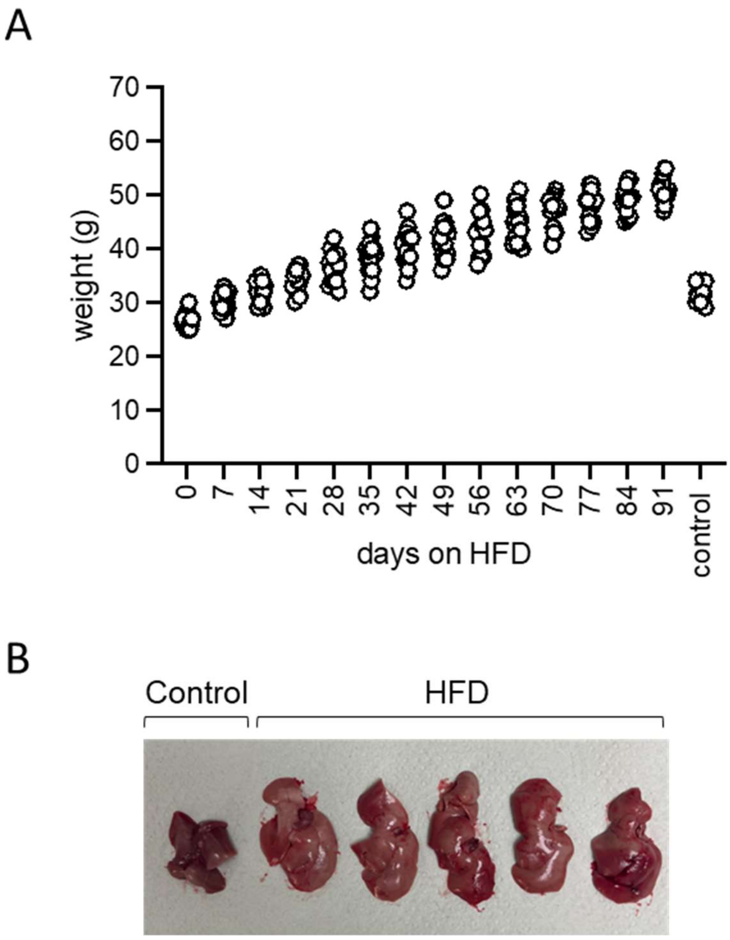


<u>**Supplementary Figure S3: Effect of a high fat diet on body weight and liver size of mice**</u>

C57BL/6 mice were maintained on a HFD or kept on a normal diet as controls for 13 weeks prior to reovirus treatment. **(A)** Mice weights were recorded weekly (n=7 per group). **(B)** Photographs of an example control liver compared to HFD livers.
