## Supplementary Figure 4 for "Single versus repeated intravenous oncolytic reovirus infusions: Implications for immune modulation and rationalised scheduling of therapy in hepatocellular carcinoma"

**A**

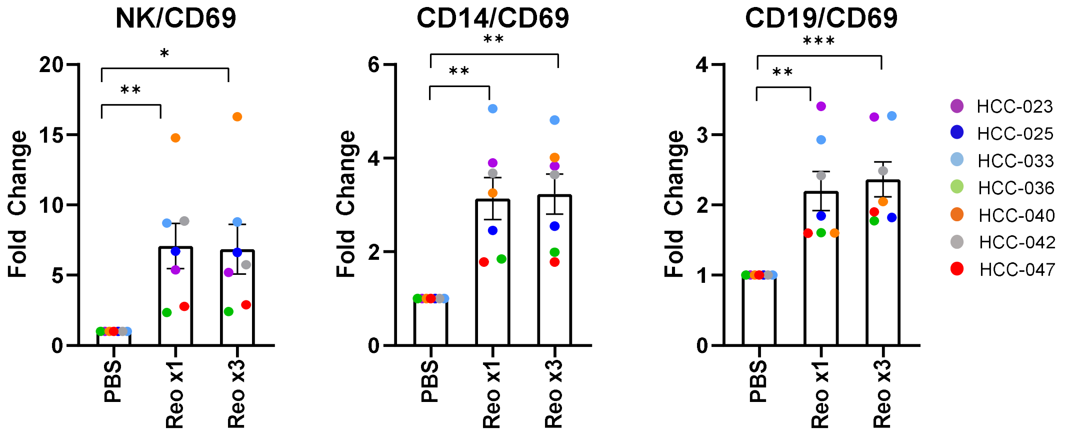


**B**

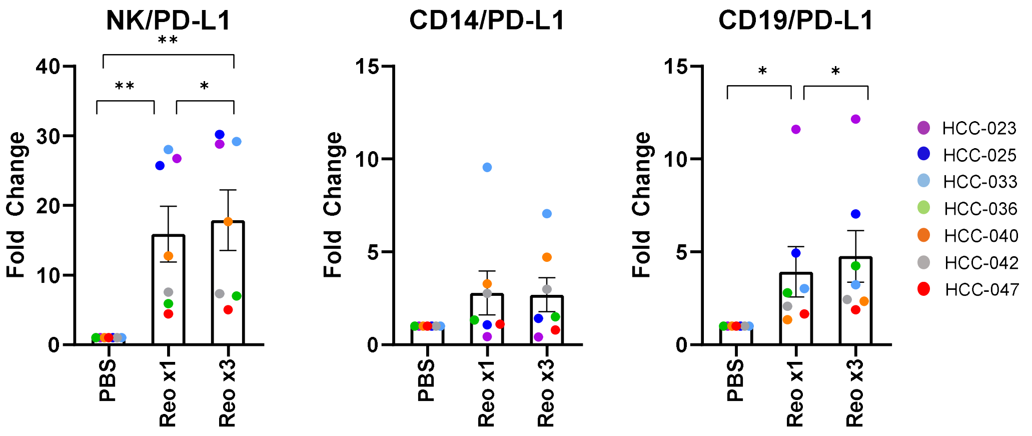


**Supplementary Figure S4: Immunophenotyping of HCC patient PBMC in response to single or repeated doses of reovirus**

PBMCs were isolated from the blood of patients with HCC. Cells were treated with PBS, one (Reox1) or repeated (Reox3) doses of reovirus. After 24 hours of culture, immunophenotyping for cell-surface **(A)** CD69 and **(B)** PD-L1 was performed. Data is presented as fold change (± SEM) from PBS sample; *P<0.05, **P<0.01, ***P<0.001; n=7.
