## Supplementary Figure 5 for "Single versus repeated intravenous oncolytic reovirus infusions: Implications for immune modulation and rationalised scheduling of therapy in hepatocellular carcinoma"

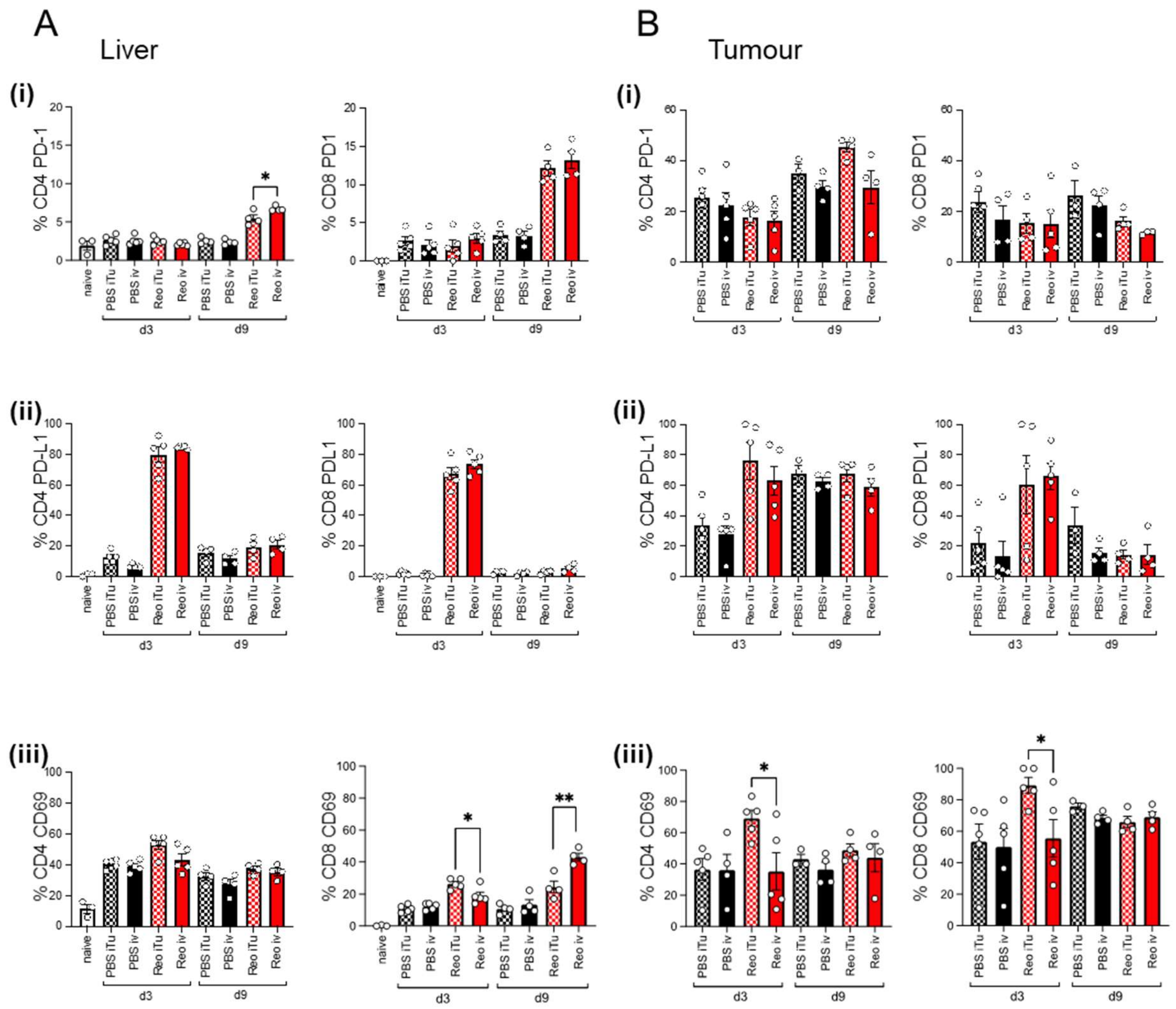


**Supplementary Figure S5: Effect of a single dose of *i.v.* vs *i.t.* reovirus on PD-1, PD-L1 and CD69 expression by organ-resident T cells**

1MEA tumour-bearing mice were treated with a single dose of *i.v.* or *i.t.* reovirus or PBS (naïve represents untreated mice) prior to organ harvest at 3 or 9 days post-treatment. Single cell suspensions of liver (A) and tumour (B) were analysed by flow cytometry for (i) PD-1, (ii) PD-L1 and (iii) CD69 expression on CD4+ and CD8+ T cells. Data is presented as mean % positive ± SEM for n=5 mice per group for naïve (white bars), PBS (black bars) and reovirus (red bars) for *i.v.* reovirus (solid bars) or *i.t.* reovirus (hatched bars); *P<0.05, **P<0.01.
