## Supplementary Figure 6 for "Single versus repeated intravenous oncolytic reovirus infusions: Implications for immune modulation and rationalised scheduling of therapy in hepatocellular carcinoma"

**A**

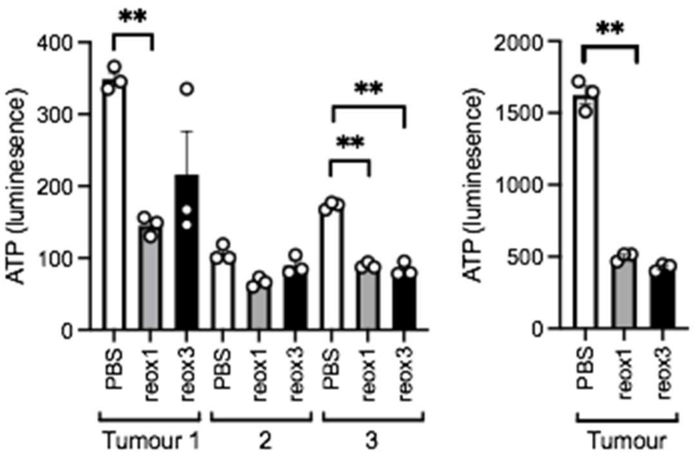


**B**

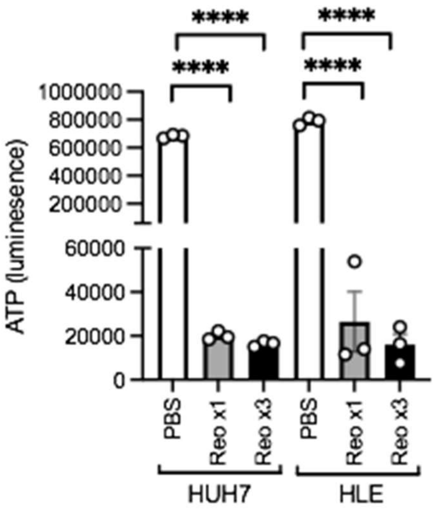


<u>**Supplementary Figure S6: Viability of HCC tumour and HCC cell lines in response to single or repeated doses of reovirus**</u>

**(A)** Single-cell suspensions of HCC tumour and **(B)** HCC cell lines were treated with PBS (white bars), one (Reox1; grey bars) or repeated doses (Reox3; black bars) of reovirus. After 24 hours of culture, ATP levels within cultures were measured. Data is presented as luminescence ± SEM from n=3 HCC tumours from one individual patient and n=1 tumour from a different patient; **P<0.01, ****P<0.0001.
