## Supplementary Table 3 for "Single versus repeated intravenous oncolytic reovirus infusions: Implications for immune modulation and rationalised scheduling of therapy in hepatocellular carcinoma"

Supplementary Table 3: Raw data values for immunophenotyping analyses in **(A)** repeated dose and **(B)** single dose patients.

**Suppl. Table 3A**

**REPEATED DOSE TRIAL**

| **CD4/CD69** | **day 1 pre** | **day 1 post** | **day 3** | **day 5** | **pre-surgery** | **1M** |
|---|---|---|---|---|---|---|
| | 3.80 | 4.23 | 8.01 | 8.29 | 7.52 | 5.47 |
| | 3.81 | 4.64 | | 6.96 | 5.45 | |
| | 4.33 | 15.53 | 10.15 | 9.36 | 4.18 | 4.98 |
| | 4.82 | 5.86 | 7.99 | 5.25 | 4.32 | 4.07 |

| **CD8/CD69** | **day 1 pre** | **day 1 post** | **day 3** | **day 5** | **pre-surgery** | **1M** |
|---|---|---|---|---|---|---|
| | 4.61 | 5.45 | 8.99 | 10.74 | 7.30 | 8.21 |
| | 4.14 | 4.24 | | 11.90 | 7.52 | |
| | 5.86 | 19.16 | 17.18 | 19.72 | 19.83 | 10.69 |
| | 4.75 | 4.60 | 10.23 | 8.92 | 6.35 | 3.77 |

| **CD14/CD69** | **day 1 pre** | **day 1 post** | **day 3** | **day 5** | **pre-surgery** | **1M** |
|---|---|---|---|---|---|---|
| | 3.42 | 3.04 | 3.18 | 4.60 | 3.13 | 3.61 |
| | 3.69 | 7.65 | | 3.55 | 4.21 | |
| | 2.31 | 7.28 | 3.51 | 3.29 | 4.67 | 4.30 |
| | 6.70 | 6.04 | 4.79 | 4.36 | 4.53 | 2.77 |

| **CD19/CD69** | **day 1 pre** | **day 1 post** | **day 3** | **day 5** | **pre-surgery** | **1M** |
|---|---|---|---|---|---|---|
| | 3.27 | 4.92 | 5.18 | 5.26 | 7.84 | 4.00 |
| | 3.70 | 3.60 | | 3.46 | 2.41 | |
| | 2.77 | 17.08 | 4.52 | 3.31 | 4.79 | 4.96 |
| | 6.60 | 3.02 | 3.13 | 2.95 | 3.53 | 2.80 |

**Suppl. Table 3B**

**SINGLE DOSE TRIAL**

| **CD4/CD69** | **day 1 pre** | **day 1 mid** | **day 1 post** | **day 3** | **pre-surgery** | **1M** |
|---|---|---|---|---|---|---|
| | 5.9 | 4.0 | 4.7 | 8.2 | 7.4 | 4.7 |
| | 4.6 | 4.7 | 4.1 | | 5.7 | 4.3 |
| | 4.7 | 3.5 | 3.3 | 11.7 | 6.9 | 4.1 |
| | 4.2 | 3.2 | 3.3 | 10.0 | 5.8 | 3.6 |
| | 3.6 | 3.6 | 4.0 | 13.0 | 5.1 | 4.8 |
| | 5.0 | 4.8 | 5.4 | 12.8 | 6.0 | 4.4 |
| | 4.1 | 5.0 | 4.9 | 10.1 | | 4.2 |
| | 4.6 | 4.5 | 4.0 | 9.2 | 4.0 | 4.1 |
| | 4.8 | 4.5 | 4.0 | 6.0 | 3.3 | 4.5 |

| **CD8/CD69** | **day 1 pre** | **day 1 mid** | **day 1 post** | **day 3** | **pre-surgery** | **1M** |
|---|---|---|---|---|---|---|
| | 6.3 | 3.5 | 5.6 | 10.0 | 7.6 | 6.0 |
| | 4.4 | 5.8 | 5.0 | | 7.5 | 5.0 |
| | 4.8 | 3.4 | 3.2 | 11.5 | 7.0 | 4.4 |
| | 3.9 | 3.3 | 3.3 | 12.6 | 7.8 | 4.1 |
| | 4.3 | 4.8 | 4.3 | 12.7 | 6.6 | 6.0 |
| | 5.1 | 5.6 | 5.8 | 9.1 | | 6.6 |
| | 5.2 | 5.5 | 4.8 | 16.1 | 4.7 | 4.6 |
| | 4.1 | 4.6 | 4.3 | 5.7 | 3.5 | 5.4 |

| **CD14/CD69** | **day 1 pre** | **day 1 mid** | **day 1 post** | **day 3** | **pre-surgery** | **1M** |
|---|---|---|---|---|---|---|
| | 3.9 | 3.7 | 4.1 | 4.4 | 4.1 | 5.5 |
| | 3.6 | 3.4 | 4.0 | | 5.1 | 3.6 |
| | 3.7 | 4.4 | 2.9 | 4.7 | 3.1 | 2.9 |
| | 3.8 | 3.0 | 3.2 | 3.3 | 3.1 | 3.5 |
| | 4.0 | 3.7 | 3.4 | 5.3 | 4.1 | 3.5 |
| | 3.4 | 4.0 | 4.7 | 5.5 | 4.2 | 3.3 |
| | 4.9 | 3.9 | 4.2 | 4.7 | | 4.5 |
| | 4.5 | 4.0 | 3.7 | 6.1 | 3.8 | 4.0 |
| | 4.1 | 4.3 | 3.5 | 4.9 | 5.1 | 3.3 |

| **CD19/CD69** | **day 1 pre** | **day 1 mid** | **day 1 post** | **day 3** | **pre-surgery** | **1M** |
|---|---|---|---|---|---|---|
| | 5.0 | 4.6 | 4.5 | 4.4 | 5.9 | 5.3 |
| | 3.7 | 4.5 | 4.2 | | 5.2 | 5.9 |
| | 5.7 | 4.9 | 4.3 | 12.9 | 5.6 | 4.7 |
| | 4.9 | 4.3 | 4.2 | 9.1 | 4.4 | 4.0 |
| | 5.2 | 4.9 | 5.1 | 13.3 | 5.9 | 5.3 |
| | 6.0 | 7.3 | 6.6 | 14.9 | 8.5 | 5.3 |
| | 6.3 | 6.2 | 7.2 | 15.0 | | 5.9 |
| | 8.2 | 6.7 | 5.8 | 8.9 | 5.6 | 6.7 |
| | 8.2 | 7.1 | 7.1 | 10.0 | 10.7 | 4.9 |
