## Supplementary Table 4 for "Single versus repeated intravenous oncolytic reovirus infusions: Implications for immune modulation and rationalised scheduling of therapy in hepatocellular carcinoma"

Supplementary Table 4: Raw data values for chemokine analyses for (A) repeated dose and (B) single dose patients.

**Suppl. Table 4A**

| REPEATED DOSES TRIAL | | | | |
|---|---|---|---|---|
| | **day 1 pre** | **day3** | **day5** | **pre-surgery** |
| **IP-10** | 124.44 | 570.88 | 583.71 | 137.1 |
| | 279.13 | 2199.51 | 1158.67 | 237.73 |
| | 149.66 | 1774.07 | 652.4 | 247.93 |
| | 162.61 | 2051.77 | 701.84 | 455.52 |
| | 195.65 | 2283.24 | 641.1 | 196.77 |
| | 872.21 | 1776.6 | 891.52 | 269.95 |
| | **day 1 pre** | **day3** | **day5** | **pre-surgery** |
| **MIG** | 82.95 | 177.28 | 360.34 | 104.14 |
| | 246.63 | 808.23 | 551.29 | 248.62 |
| | 170.81 | 400.7 | 369.04 | 241.04 |
| | 164.73 | 391.56 | 360.47 | 253.54 |
| | 125.93 | 755.75 | 447 | 192.86 |
| | 1665.47 | 731.19 | 464.78 | 149.53 |
| | **day 1 pre** | **day3** | **day5** | **pre-surgery** |
| **MIP-1b** | 25.4 | 24.71 | 25.92 | 23.23 |
| | 28.97 | 39.87 | 27.19 | 19.84 |
| | 24.46 | 45.37 | 23.29 | 30.39 |
| | 30.68 | 32.35 | 22.57 | 23.4 |
| | 17.11 | 33.33 | 21 | 20.33 |
| | 65.66 | 31.39 | 27.09 | 25.34 |

**Suppl. Table 4B**

| SINGLE DOSE TRIAL | | | | |
|---|---|---|---|---|
| | **day 1 pre** | **day 1 post** | **day 3** | **pre-surgery** |
| **IP-10** | 78.68 | 58.6 | 1156.35 | 284.52 |
| | 143.9 | 174.54 | | 648.69 |
| | 497.36 | 488.59 | 11615.45 | 2319.11 |
| | 194.19 | 178.61 | 3124.89 | 1773.06 |
| | 374 | 257.59 | 3370.07 | |
| | 201.9 | 170.31 | 3004.39 | 472 |
| | 685.81 | 621.69 | 3031.21 | |
| | 210.13 | 276.11 | 3739.2 | 297.21 |
| | 244.47 | 198.88 | 1147.82 | 235.27 |
| | **day 1 pre** | **day 1 post** | **day 3** | **pre-surgery** |
| **MIG** | 111.35 | 85.88 | 415.85 | 139.58 |
| | 342.71 | 354.67 | | 534.58 |
| | 443.24 | 585.56 | 2517.59 | 1243.83 |
| | 236.46 | 314.56 | 2365.42 | 1796.11 |
| | 265.38 | 235.14 | 692.22 | |
| | 224.6 | 208.22 | 1825.76 | 782.92 |
| | 413.78 | 399.29 | 732.91 | |
| | 296.54 | 623.9 | 1564.51 | 212.72 |
| | 200.82 | 170.82 | 347.91 | 261.96 |
| | **day 1 pre** | **day 1 post** | **day 3** | **pre-surgery** |
| **MIP-1b** | 35.06 | 24.94 | 58.44 | 40.79 |
| | 66.3 | 65.41 | | 57.99 |
| | 49.58 | 46.79 | 111.71 | 48.27 |
| | 37.63 | 35.39 | 56.63 | 36.81 |
| | 42.67 | 32.54 | 87.58 | |
| | 35.24 | 31.25 | 83.52 | 22.73 |
| | 99.34 | 85.59 | 103.09 | |
| | 49.5 | 44.44 | 84.34 | 35.77 |
| | 60.66 | 42.13 | 65.08 | 58.12 |
