## Supplementary Table 1 for "Single versus repeated intravenous oncolytic reovirus infusions: Implications for immune modulation and rationalised scheduling of therapy in hepatocellular carcinoma"

### Suppl. Table 1A

| REPEATED DOSE TRIAL | | |
|---|---|---|
| **Age** | **Sex** | **Previous Therapy** |
| 70s | M | Nil |
| 60s | M | Nil |
| 70s | M | Capecitabine + radiotherapy |
| 60s | M | Nil |
| 60s | M | Nil |
| 50s | F | Oxaliplatin |
| 50s | M | Nil |
| 60s | F | Nil |
| 70s | M | Nil |
| 60s | M | Nil |

### Suppl. Table 1B

| SINGLE DOSE TRIAL | | | |
|---|---|---|---|
| **Age** | **Sex** | **Previous Therapy** | **Steroid Dose** |
| 50s | M | Surgery + TMZ Chemoradiotherapy | 8 mg |
| 70s | M | Oxaliplatin + radiotherapy | 2 mg |
| 40s | F | Surgery + radiotherapy<br>PCV<br>Surgery | None |
| 40s | F | Surgery + radiotherapy<br>PCV<br>TMZ | None |
| 60s | M | Nil | 2 mg |
| 60s | M | Surgery + TMZ Chemoradiotherapy | 2 mg |
| 60s | M | Surgery + TMZ Chemoradiotherapy<br>CCNU<br>Bevacizamub | None |
| 60s | F | Surgery + TMZ Chemoradiotherapy | 6 mg |
| 70s | F | Surgery | 8 mg |

TMZ: temozolomide; PCV: procarbazine; CCNU: lomustine

Supplementary Table 1: Patient demographics for **(A)** Repeated dose trial and **(B)** Single dose trial.
