## Supplementary Table 2 for "Single versus repeated intravenous oncolytic reovirus infusions: Implications for immune modulation and rationalised scheduling of therapy in hepatocellular carcinoma"

Supplementary Table 2: Raw values (pg/ml) for cytokine analyses from **(A)** repeated dose and **(B)** single dose patients.

**Suppl. Table 2A**

**REPEATED DOSE TRIAL**

| | day 1 pre | day3 | day5 | pre-surgery |
|---|---|---|---|---|
| **IFN-a** | 40.24 | 50.35 | 38.81 | 46.68 |
| | 33.43 | 36.83 | 39.53 | 34.51 |
| | 33.43 | 41.86 | 35.29 | 31.49 |
| | 34.24 | 41.17 | 38.81 | 41.86 |
| | 33.97 | 50.35 | 50.35 | 43.66 |
| | 54.01 | 50.94 | 36.07 | 31.2 |

| | day 1 pre | day3 | day5 | pre-surgery |
|---|---|---|---|---|
| **IL2-Ra** | 54.82 | 91.39 | 73.27 | 59.96 |
| | 59.96 | 81.4 | 75.98 | 62.72 |
| | 56.01 | 84.09 | 66.25 | 63.9 |
| | 31.73 | 51.64 | 42.8 | 50.44 |
| | 50.44 | 104.73 | 85.25 | 66.25 |
| | 91.39 | 113.82 | 81.4 | 45.63 |

| | day 1 pre | day3 | day5 | pre-surgery |
|---|---|---|---|---|
| **IL-16** | 233.74 | 303.91 | 285.44 | 294.75 |
| | 377.21 | 415.38 | 430.98 | 375.15 |
| | 354.29 | 455.82 | 358.51 | 339.34 |
| | 310.7 | 390.46 | 339.34 | 297.05 |
| | 237.61 | 375.15 | 361.66 | 394.49 |
| | 387.42 | 401.51 | 280.72 | 332.84 |

| | day 1 pre | day3 | day5 | pre-surgery |
|---|---|---|---|---|
| **IL-18** | 10.65 | 19.9 | 42.09 | 12.28 |
| | 20.61 | 24.14 | 26.11 | 20.35 |
| | 31.23 | 52.11 | 67.51 | 37.64 |
| | 26.8 | 27.7 | 38.71 | 28.79 |
| | 1.52 | 8.1 | 10.74 | 10.57 |
| | 27.92 | 42.59 | 41.77 | 22.97 |

| | day 1 pre | day3 | day5 | pre-surgery |
|---|---|---|---|---|
| **IL-4** | 4.32 | 3.08 | 2.99 | 3.05 |
| | 0.54 | 1.1 | 0.88 | 0.7 |
| | 2.69 | 2.17 | 1.8 | 1.67 |
| | 1.67 | 0.9 | 1.44 | 1.75 |
| | 3.19 | 1.97 | 1.99 | 2.3 |
| | 2.28 | 1.57 | 2.45 | 1.99 |

| | day 1 pre | day3 | day5 | pre-surgery |
|---|---|---|---|---|
| **IL-5** | 16.02 | 11.49 | 10.45 | 9.8 |
| | 0.49 | | 2.35 | 3.19 |
| | 3.39 | 2.53 | 3.87 | 2.53 |
| | 5.46 | 1.75 | 3.39 | 5.6 |
| | 9.58 | 4.02 | 6.39 | 6.77 |
| | 11.08 | 8.1 | 28.21 | 8.79 |

**Suppl. Table 2B**

**SINGLE DOSE TRIAL**

| | day 1 pre | day 1 post | day 3 | pre-surgery |
|---|---|---|---|---|
| **IFN-a** | 32.89 | 33.34 | 41.03 | 26.69 |
| | 34.46 | 33.79 | | 38.47 |
| | 37.86 | 37.03 | 60.87 | 50.72 |
| | 31.49 | 32.19 | 44.02 | 46.68 |
| | 30.53 | 27.76 | 37.03 | |
| | 26.15 | 24.45 | 40.25 | 39.47 |
| | 40.25 | 35.76 | 53.12 | |
| | 27.23 | 33.11 | 43.65 | 28.28 |
| | 17.93 | 22.03 | 24.45 | 23.27 |

| | day 1 pre | day 1 post | day 3 | pre-surgery |
|---|---|---|---|---|
| **IL2-Ra** | 54.58 | 52.83 | 70.59 | 38.67 |
| | 99.45 | 107.41 | | 112 |
| | 121.57 | 143.46 | 196.06 | 175.77 |
| | 76.59 | 87.64 | 177.4 | 237.16 |
| | 43.13 | 46.67 | 52.83 | |
| | 63.27 | 71.02 | 109.08 | 104.9 |
| | 101.54 | 101.54 | 149.21 | |
| | 44.02 | 71.88 | 89.76 | 54.58 |
| | 60.67 | 75.73 | 82.55 | 67.58 |

| | day 1 pre | day 1 post | day 3 | pre-surgery |
|---|---|---|---|---|
| **IL-16** | 112.13 | 127.03 | 158.58 | 88.88 |
| | 157.18 | 157.18 | | 174.53 |
| | 222.62 | 242 | 267.35 | 272.36 |
| | 115.14 | 154.37 | 181.36 | 196.88 |
| | 145.86 | 126.3 | 146.57 | |
| | 90.47 | 113.64 | 137.96 | 130.7 |
| | 137.24 | 135.79 | 180.68 | |
| | 95.2 | 177.27 | 202.21 | 119.63 |
| | 110.61 | 143 | 184.76 | 144.43 |

| | day 1 pre | day 1 post | day 3 | pre-surgery |
|---|---|---|---|---|
| **IL-18** | 41.38 | 43.13 | 66.5 | 40.64 |
| | 54.48 | 79.85 | | 82.65 |
| | 33.15 | 40.27 | 59.28 | 68.81 |
| | 44.7 | 54.11 | 81.44 | 105.68 |
| | 26.47 | 26.75 | 44.06 | |
| | 16.13 | 19.06 | 32.5 | 36.58 |
| | 43.04 | 44.89 | 78.37 | |
| | 36.48 | 51.81 | 89.64 | 47.29 |
| | 39.53 | 45.99 | 71.5 | 64.09 |

| | day 1 pre | day 1 post | day 3 | pre-surgery |
|---|---|---|---|---|
| **IL-4** | 4.22 | 3.04 | 2.4 | 4.39 |
| | 5.72 | 4.15 | | 4.32 |
| | 5.17 | 0.27 | 0.79 | 1.13 |
| | 6.5 | 3.76 | 3.27 | 0.67 |
| | 1.82 | 1.08 | 0.23 | |
| | 3.9 | 1.78 | 1.59 | 1.39 |
| | 4.32 | 4.19 | 3 | |
| | 7.54 | 2.68 | 2.77 | 5.14 |
| | 6.16 | 4.11 | 4.11 | 4.19 |

| | day 1 pre | day 1 post | day 3 | pre-surgery |
|---|---|---|---|---|
| **IL-5** | 29.84 | 25.12 | 20.6 | 29.84 |
| | 46.26 | 33.34 | | 44.33 |
| | 41.61 | 26.42 | 20.6 | 23.44 |
| | 64.86 | 49.07 | 47.67 | 21.69 |
| | 24.79 | 20.6 | 16.31 | |
| | 51.57 | 41.86 | 31.62 | 22.75 |
| | 52.24 | 55.75 | 44.57 | |
| | 52.47 | 30.44 | 30.74 | 39.56 |
| | 56.83 | 52.47 | 41.1 | 44.82 |
